# Environmental Chemical Exposures and Accelerated Epigenetic Aging: Links to Metabolic and Age-Related Health Outcomes

**DOI:** 10.64898/2026.11.10.26367792

**Authors:** Hyeongyu Shin, Jeeyoung Kim, Euikyoung Song, Nam-Eun Kim, Min Heo, Youngwoong Kim, Jeongmin Oh, Yong Min Cho, Young-Seoub Hong, Woo Jin Kim, Sungho Won

**Affiliations:** Interdisciplinary Program of Bioinformatics, College of Natural Sciences, Seoul National University, Seoul, South Korea; Department of Internal Medicine and Environmental Health Center, Kangwon National University, Chuncheon, South Korea; Department of Public Health Sciences, Seoul National University, Seoul, South Korea; Institute of Health and Environment, Seoul National University, Seoul, South Korea; Department of Nano, Chemical and Biological Engineering, SeoKyeong University, Seoul, South Korea; Department of Preventive Medicine, College of Medicine, Dong-A University, Busan, South Korea

**Author notes:** Corresponding Author: Sungho Won, Department of Public Health Sciences, Seoul National University, Seoul 08826, South Korea, Woo Jin Kim, Department of Internal Medicine and Environmental Health Center, School of Medicine, Kangwon National University, Chuncheon 24341, South Korea.

**Keywords:** PAHs, DNA methylation, GrimAge, Mediation, Mendelian randomization, Epigenetic age acceleration, Environmental mixture, Environmentally vulnerable areas

## Abstract

Environmental chemical exposures may influence biological aging, but their relationships with epigenetic aging and related health outcomes remain incompletely understood. In this study, we evaluated the associations of 13 environmental exposures, including heavy metals, polycyclic aromatic hydrocarbons (PAHs), and volatile organic compounds (VOCs), with epigenetic aging in 666 older adults from the Forensic Research via Omics Markers in Environmental Health Vulnerable Areas (FROM) cohort. Among the evaluated epigenetic aging biomarkers, GrimAge acceleration showed the most prominent associations, with 2-hydroxyfluorene (2-HF), 2-naphthol (2-NAP), blood cadmium (bCd), and methylhippuric acid (MHA) exhibiting significant positive associations (strongest for 2-HF: β = 0.491 years per doubling of exposure, *P* = 1.22 × 10□¹□). Quantile g-computation showed that combined environmental exposures were significantly associated with increased GrimAge acceleration (ψ = 0.497, 95% CI: 0.013–0.981, *P* = 0.044), and Bayesian kernel machine regression identified 2-HF and 2-NAP as major contributors. Among clinical traits, white blood cell count (WBC) and gamma-glutamyl transferase (GGT) were significantly and positively associated with GrimAge acceleration, and two-sample Mendelian randomization provided nominal support for a positive effect of GrimAge acceleration on GGT. Mediation analyses identified significant indirect effects through GrimAge acceleration for the associations of 2-HF, 2-NAP, bCd, and MHA with GGT. Exploratory disease analyses further suggested potential associations with diabetic nephropathy and osteoporosis, together with an inverse association with cardiac valvular disease. These findings suggest that environmental chemical exposures are associated with accelerated epigenetic aging, which may represent a biological pathway linking environmental exposures to metabolic and age-related health outcomes.

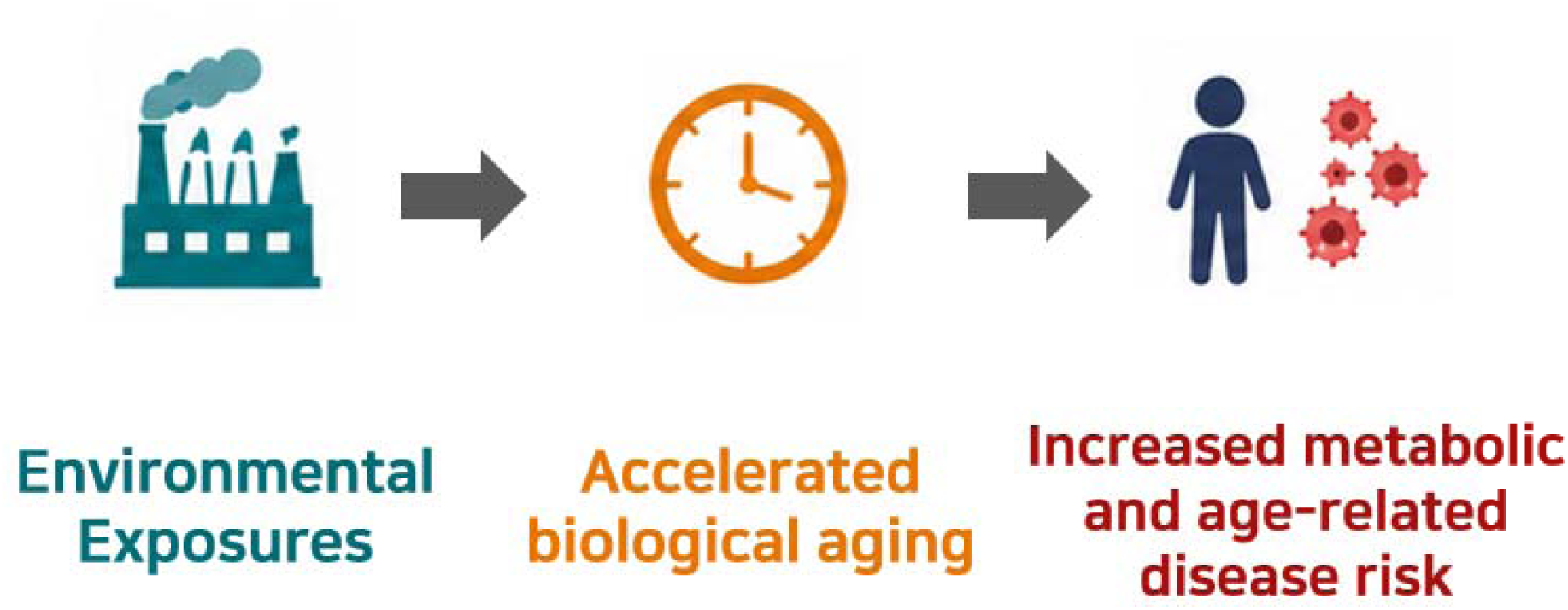

**Highlights:**

- Environmental chemical exposures were associated with accelerated GrimAge.
- PAH biomarkers 2-HF and 2-NAP were key contributors to the exposure mixture.
- GrimAge acceleration was positively associated with WBC and GGT.
- GrimAge acceleration mediated environmental exposure–GGT associations.
- GrimAge showed suggestive links to diabetic nephropathy and osteoporosis.

## 1. Introduction

Chronic exposure to environmental chemicals can induce physiological and molecular alterations that may contribute to the development and progression of chronic diseases (Landrigan et al., 2018). Heavy metals, polycyclic aromatic hydrocarbons (PAHs), and volatile organic compounds (VOCs) are major environmental hazards to which humans are widely exposed through industrial activities and the surrounding environment, and have been associated with biological processes including oxidative stress, inflammation, and metabolic dysfunction (Bolden et al., 2015; Kim et al., 2013; Tchounwou et al., 2012; Valko et al., 2005). Emerging geroscience perspectives further identify the environment as a potentially modifiable determinant of biological aging trajectories, with implications for preventing aging-related disease and reducing disparities in healthy aging (Belsky & Baccarelli, 2023). However, how these environmental exposures are associated with biological aging and how such alterations relate to human health remain incompletely understood.

DNA methylation-based epigenetic clocks have emerged as important biomarkers for quantifying biological aging (Bell et al., 2019). The Hannum and Horvath clocks were primarily developed to predict chronological age, whereas subsequent generations, including PhenoAge and GrimAge, were designed to better capture biological characteristics related to health and mortality (Hannum et al., 2013; Horvath, 2013; Levine et al., 2018; Lu et al., 2019). Epigenetic age acceleration (EAA), representing an epigenetic age greater than expected based on chronological age, may reflect accelerated biological aging. EAA may therefore serve as a potential biomarker for evaluating biological alterations associated with environmental exposures and their health implications (Fransquet et al., 2019; Hillary et al., 2020; Oblak et al., 2021).

Growing evidence suggests that exposure to air pollution, metals, and other environmental chemicals may be associated with EAA (Li et al., 2018; Nwanaji-Enwerem et al., 2020; Van der Laan et al., 2022; Ward-Caviness et al., 2016). Recent studies have further linked internal chemical exposures, including individual metals and metal mixtures, to multiple epigenetic aging measures (Boyer et al., 2023; Khodasevich et al., 2025; Mi et al., 2025). Environmental chemical exposures have also been associated with clinical alterations relevant to aging and disease, including associations between PAH metabolites and serum gamma-glutamyl transferase (GGT) (Ryu & Hong, 2024). However, these relationships have largely been investigated separately, and relatively little is known about how exposure-related alterations in epigenetic aging relate to physiological and clinical changes within the same population. Evidence also remains limited for some environmental chemicals, including VOC metabolites, in relation to DNA methylation-based aging measures. Integrating environmental exposures, epigenetic aging, and health-related phenotypes within a single cohort may therefore provide a more comprehensive understanding of the potential biological pathways linking environmental exposures to adverse health outcomes.

The FROM cohort provides an opportunity to address these gaps in older adults residing in environmentally vulnerable areas. Previous analyses of this cohort have identified heavy metal-associated multi-omic alterations and plasma proteomic profiles implicating immune, lipid, and oxidative-stress pathways (Heo et al., 2026; Kim et al., 2025). In the present study, we evaluated associations of 13 environmental exposure biomarkers, including heavy metals, PAHs, and VOCs, with multiple epigenetic aging measures and additionally assessed the effects of combined exposures. We further examined associations between epigenetic aging and clinical traits and evaluated the potential mediating role of epigenetic aging in the relationships between environmental exposures and clinical alterations. Finally, we used Mendelian randomization (MR) based on East Asian genetic instruments to investigate potential causal relationships between epigenetic aging and clinical traits and extended these analyses to metabolic and age-related disease outcomes in Biobank Japan (BBJ) (Ishigaki et al., 2020). Together, these analyses provide an integrated framework for examining potential links among environmental chemical exposures, epigenetic aging, clinical alterations, and age-related diseases.

## 2. Materials and methods

### 2.1. Study cohorts and data sources

The FROM cohort served as the primary study population (Kwon et al., 2024). The FROM study has enrolled participants in successive annual recruitment waves, and the present analyses used two non-overlapping waves recruited in 2023 and 2024. The main analyses included 666 participants from the 2024 recruitment wave (hereafter the main analytic cohort) with available DNA methylation, genotype, environmental exposure, and clinical data. Of these, 553 participants were residents of environmentally vulnerable areas, including areas affected by a waste incinerator, a coal-fired power plant, a cement factory, an area with a high concentration of factories, or high exposure to particulate matter. The remaining 113 participants were classified as residents of control areas. Control areas were located at least 10 km from major industrial facilities and were not designated as environmentally vulnerable areas. These areas were selected to have demographic characteristics, including age and sex distributions, comparable to those of the exposure areas (Kwon et al., 2024). This cohort was used for the main analyses evaluating associations between environmental exposures and EAA, mixture effects, associations between EAA and clinical traits, and mediation effects.

Additionally, genotype and clinical data from 491 participants in the 2023 recruitment wave (hereafter the MR outcome cohort), none of whom overlapped with the main analytic cohort, were used for MR analyses of clinical traits. For disease-related MR analyses, data from the Korean Genome and Epidemiology Study (KoGES) were additionally used (Kim et al., 2017). The present study included 1,226 KoGES participants with available DNA methylation and genotype data. Genetic association estimates for EAA derived from KoGES and the main analytic cohort were meta-analyzed to generate exposure summary statistics for the disease MR analyses. Genome-wide association study (GWAS) summary statistics for disease outcomes were obtained from BBJ, comprising approximately 170,000 participants. An overview of the available data and the specific roles of each cohort in the present study is provided in Fig. 1a.

**Fig. 1.**
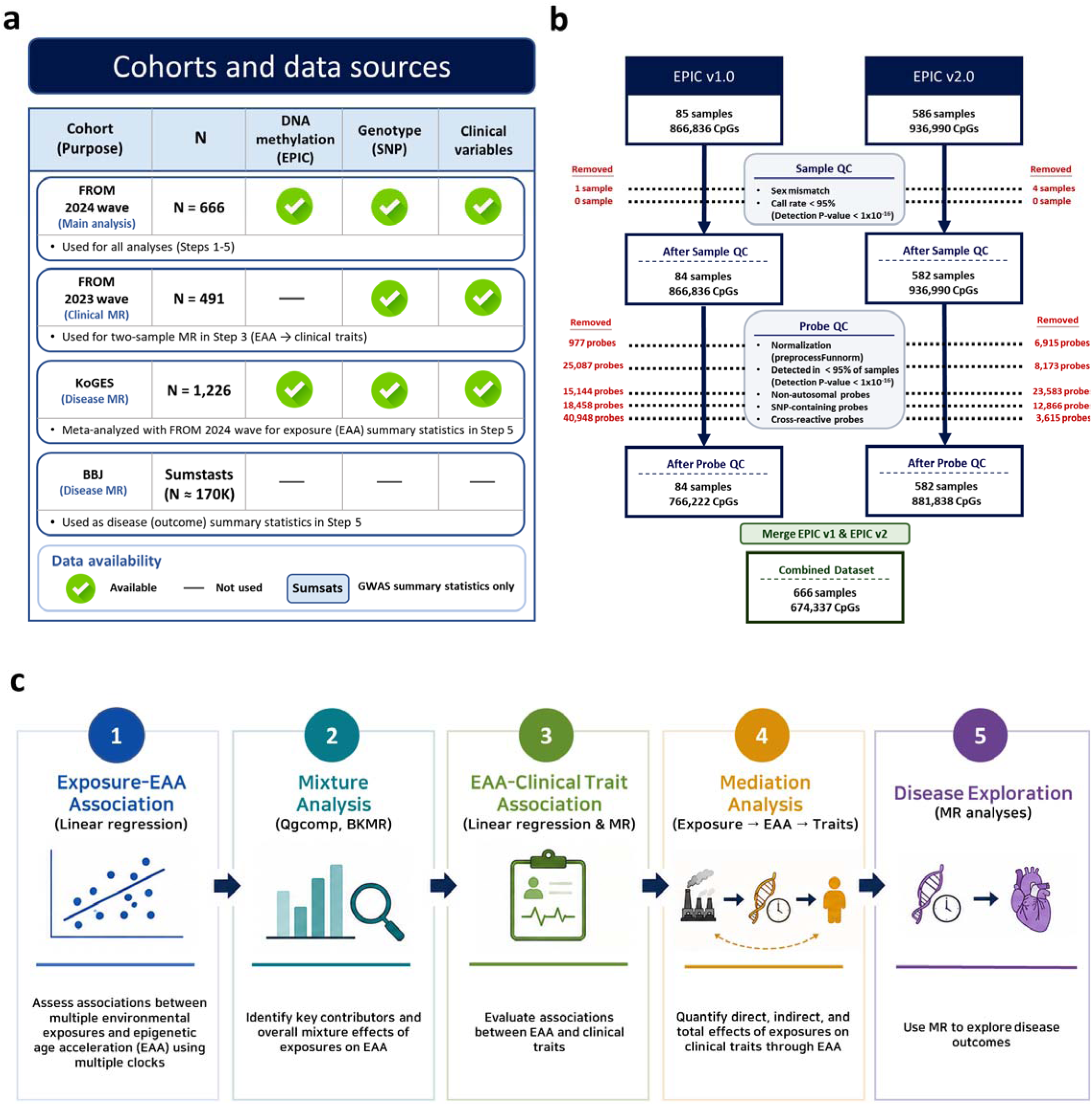
Overview of the study cohorts, DNA methylation data processing, and analytical framework. (a) Cohorts and data sources used in the study. (b) DNA methylation preprocessing and quality-control workflow for the EPIC v1.0 and v2.0 datasets. (c) Overall analytical framework comprising exposure–EAA association, mixture analysis, EAA–clinical trait analysis, mediation analysis, and disease MR.

### 2.2. Environmental exposure and clinical variable measurement

A total of 13 environmental exposure biomarkers were analyzed, including heavy metals, PAHs, and VOCs. Heavy metal-related biomarkers included blood lead (bPb), blood cadmium (bCd), blood mercury (bHg), urinary cadmium (uCd), and the sum of urinary arsenic species (As^III^, As^V^, and MMA). PAH metabolites included 2-naphthol (2-NAP), 1-hydroxypyrene (1-OHP), 2-hydroxyfluorene (2-HF), and 1-hydroxyphenanthrene (1-HPH), while VOC metabolites included phenylglyoxylic acid (PGA), trans,trans-muconic acid (tt-MA), methylhippuric acid (MHA), and benzylmercapturic acid (BMA). All urinary exposure biomarkers were adjusted for urinary creatinine concentration, whereas bPb, bCd, and bHg were analyzed as blood-based biomarkers.

A total of 24 clinical variables were evaluated: platelet count, hemoglobin, hematocrit, white blood cell count (WBC), red blood cell count (RBC), mean corpuscular volume (MCV), mean corpuscular hemoglobin (MCH), mean corpuscular hemoglobin concentration (MCHC), total protein, serum albumin, total calcium, inorganic phosphate, aspartate transaminase (AST), alanine transaminase (ALT), gamma-glutamyl transferase (GGT), alkaline phosphatase (ALP), lactic acid dehydrogenase (LDH), blood urea nitrogen (BUN), total cholesterol, triglyceride (TG), high-density lipoprotein cholesterol (HDL), low-density lipoprotein cholesterol (LDL), N-acetyl-β-D-glucosaminidase (NAG), and beta-2 microglobulin (β2-MG). β2-MG measurements were unavailable for nine participants; therefore, analyses involving β2-MG were conducted using the 657 participants with available measurements. Detailed procedures for sample collection and measurement of environmental exposure biomarkers and clinical variables have been described previously (Kwon et al., 2024). The study was approved by the Institutional Review Board of Kangwon National University Hospital (KNUH-B-2021-06-001) and conducted in accordance with the Declaration of Helsinki and relevant guidelines and regulations. Written informed consent was obtained from all participants.

### 2.3. DNA methylation profiling and preprocessing

Genomic DNA isolated from peripheral blood was used for DNA methylation profiling. Genome-wide DNA methylation was measured using the Infinium MethylationEPIC BeadChip (Illumina, San Diego, CA, USA), including data generated using the EPIC v1.0 (n = 85) and v2.0 (n = 586) arrays. DNA methylation data preprocessing and quality control were performed using the minfi R package (Aryee et al., 2014). Five subjects showing a mismatch between reported sex and sex predicted from X- and Y-chromosome probe intensities were excluded. All included samples showed detection *P* < 1 × 10□¹□ for more than 95% of probes. Background adjustment, dye-bias correction, and functional normalization were performed using the preprocessFunnorm function.

For CpG-level QC, probes with detection *P* > 1 × 10□¹□ in more than 5% of samples were excluded. Probes located on sex chromosomes, probes containing SNPs at the CpG or single-base extension sites, and known cross-reactive probes were also removed. The EPIC v2.0 probes were subsequently mapped to their corresponding v1.0 probe identifiers to harmonize the two array datasets, resulting in 666 subjects and 674,337 CpG sites for downstream analyses (Fig. 1b). To minimize technical variability, batch effects across 87 array slides were corrected using the ComBat function from the sva R package (Leek et al., 2012). Multidimensional scaling (MDS) plots were examined before and after ComBat correction, and no clear separation between EPIC v1.0 and v2.0 samples was observed after correction (Fig. S1). Blood cell-type proportions, including CD4+ T cells, CD8+ T cells, NK cells, B cells, and monocytes, were estimated using the Houseman algorithm with an IDOL-optimized blood reference panel (Houseman et al., 2012; Koestler et al., 2016).

### 2.4. Estimation of epigenetic aging measures

Seven DNA methylation-based epigenetic aging measures were estimated, including the Horvath, Hannum, Zhang, PhenoAge, GrimAge, epiTOC, and DunedinPACE measures (Belsky et al., 2022; Hannum et al., 2013; Horvath, 2013; Levine et al., 2018; Lu et al., 2019; Yang et al., 2016; Zhang et al., 2019). Horvath, Hannum, Zhang, PhenoAge, epiTOC, and DunedinPACE were calculated using the dnaMethyAge R package (Wang et al., 2024), whereas GrimAge v1 was estimated using the meffonym R package (Suderman). For CpG sites required by individual algorithms but unavailable in the preprocessed methylation dataset, reference-based imputation implemented in the corresponding algorithms was applied.

For Horvath, Hannum, Zhang, PhenoAge, and GrimAge, age acceleration (AA) was calculated to quantify epigenetic aging independent of chronological age (Horvath & Raj, 2018). Each epigenetic age estimate was regressed on chronological age using a linear regression model, and the resulting residuals were defined as AA. Positive AA values therefore indicated faster epigenetic aging than expected based on chronological age. As epiTOC and DunedinPACE reflect mitotic aging and the pace of aging, respectively, their original estimates were used in subsequent analyses without calculating AA (Fig. S2).

### 2.5. SNP genotyping and quality control

Blood-derived DNA from FROM participants was genotyped using KoreanChip version 1.1 (Moon et al., 2019). To minimize batch effects during genotype calling, a K-medoid clustering-based approach was applied (Seo et al., 2019), and genotype quality control was performed using PLINK and ONETOOL (Song et al., 2018). At the subject level, participants with sex discrepancies or genotype call rates below 95% were excluded. At the variant level, single nucleotide polymorphisms (SNPs) with call rates below 95% or Hardy–Weinberg equilibrium (HWE) *P* < 1 × 10□□ were removed.

Genotype imputation was performed using the Korean Reference Genome panel through the Korean Imputation Server (Hwang et al., 2022), with Eagle v2.4 for pre-phasing and Minimac4 for imputation. Following imputation, SNPs with R² < 0.8, duplicate variants, genotype missing rates >5%, HWE *P* < 1 × 10□□, or minor allele frequencies (MAFs) <5% were excluded. Subject-level QC was further performed using an identity-by-state threshold of 0.99, and outliers were removed based on multidimensional scaling using LD-pruned SNPs. SNP coordinates were harmonized to the human genome build hg38 using the rtracklayer R package (Lawrence et al., 2009). A total of 5,030,771 SNPs were retained for subsequent analyses.

KoGES genotype data were generated using the Affymetrix 5.0 or 6.0 Array or the Korea Biobank Array version 1.0 or 1.1. Variants were excluded if they had a missing rate >5%, HWE *P* < 1 × 10□□, or MAF <5%, while participants were excluded for sex discrepancies, missing rates >5%, or heterozygosity outliers (*P* < 1 × 10□□). Genotype imputation was performed using the Northeast Asian Reference Database imputation server, and SNPs with an INFO score >0.8 were retained for subsequent analyses. Detailed procedures for KoGES genotyping, QC, and imputation have been described previously (Seo et al., 2024).

### 2.6. Statistical analysis

#### 2.6.1. Comparison of epigenetic aging measures between exposure and control groups

Differences in epigenetic aging measures were compared between residents of environmentally vulnerable areas (exposure group) and those from control areas (control group) using Welch’s two-sample t-tests. Multiple testing was controlled using the Benjamini–Hochberg false discovery rate (BH-FDR) method (Benjamini & Hochberg, 1995). The distributions of epigenetic aging measures were presented using violin plots with embedded box plots.

#### 2.6.2. Associations between environmental exposures and epigenetic aging

As the first step of the analytical framework (Fig. 1c), associations between the 13 environmental exposure biomarkers and epigenetic aging measures were evaluated using multiple linear regression. Urinary exposure biomarkers were adjusted for urinary creatinine, and all environmental exposure variables were log2-transformed prior to analysis. Each environmental exposure biomarker was evaluated separately, with adjustment for age, sex, smoking status, BMI, and estimated blood cell proportions. Regression coefficients, standard errors, and *P* values were obtained from each model. Multiple testing was controlled using the BH-FDR method across the 13 environmental exposures separately for each epigenetic aging measure, with FDR < 0.05 considered statistically significant.

For GrimAge AA, the robustness of associations involving urinary exposure biomarkers to the method of urinary dilution adjustment was assessed in a sensitivity analysis using uncorrected urinary concentrations, with urinary creatinine included as a separate covariate instead of using creatinine-standardized concentrations. To assess potential nonlinear effects of chronological age, a sensitivity analysis for GrimAge AA was performed by replacing the linear age term in the exposure–GrimAge AA regression models with a natural cubic spline with 3 degrees of freedom, while retaining the same exposure variables and other covariates as in the primary models. To further assess potential residual confounding by smoking, additional sensitivity analyses for GrimAge AA were performed by replacing smoking status with cumulative smoking exposure quantified as pack-years and by restricting the analysis to never-smokers. Pack-years were calculated as the number of cigarettes smoked per day divided by 20 and multiplied by the duration of smoking in years, with never-smokers assigned a value of zero. The same exposure variables and other covariates as in the primary models were retained. For each sensitivity analysis, multiple testing across the 13 environmental exposures was controlled using the BH-FDR method. Additional sensitivity analyses for GrimAge AA were conducted by stratifying the study population by sex and age (<70 and ≥70 years), with 70 years selected as the cutoff because it approximated the mean age of the study population (69.76 years). Effect modification was further assessed in multivariable regression models by including exposure-by-sex interaction terms, with men as the reference group, and exposure-by-age interaction terms, with age modeled as a continuous variable.

#### 2.6.3. Environmental mixture analysis

Following the individual exposure analyses (Fig. 1c), quantile g-computation (qgcomp) (Keil et al., 2020) and Bayesian kernel machine regression (BKMR) (Bobb et al., 2015) were performed to evaluate the association between combined environmental exposures and GrimAge AA. The mixture comprised 12 environmental exposure biomarkers, including heavy metals, PAHs, and VOCs. Because of the correlation between blood and urinary Cd (Spearman ρ = [0.552]), blood Cd was excluded from the mixture analysis to minimize redundancy, and urinary Cd was used as the representative measure of Cd exposure. All models were adjusted for age, sex, smoking status, BMI, and estimated blood cell proportions (CD8+ T cells, CD4+ T cells, NK cells, B cells, and monocytes). To assess the robustness of the mixture analysis results to the choice of Cd biomarker, sensitivity analyses were performed by replacing urinary Cd with blood Cd in both the qgcomp and BKMR models, while retaining all other exposure variables, covariates, and model specifications.

Qgcomp was performed using the qgcomp R package (Keil et al., 2020). Environmental exposures were categorized into quartiles (q = 4), and the overall mixture effect was estimated as the change in GrimAge AA associated with a simultaneous one-quartile increase in all mixture components. An additive linear model was used to estimate the overall mixture effect and the relative contribution (weight) of each exposure in the positive or negative direction. A marginal structural model with 1,000 bootstrap iterations was additionally used to estimate the overall mixture effect and its 95% confidence interval (CI).

BKMR was performed using the bkmr R package to evaluate the overall mixture effect and potential nonlinear exposure–response relationships of individual exposures (Bobb et al., 2015). Environmental exposure variables were standardized prior to analysis, and 10,000 Markov chain Monte Carlo iterations were performed with variable selection. The relative importance of individual exposures within the mixture was evaluated using posterior inclusion probabilities (PIPs). Exposure–response relationships between individual exposures and GrimAge AA were further evaluated while holding the remaining mixture components at their median levels.

#### 2.6.4. Associations and causal inference between epigenetic aging and clinical traits

As the next step of the analytical framework (Fig. 1c), associations between GrimAge AA and 24 clinical traits were evaluated using multiple linear regression. All clinical traits were log2-transformed prior to analysis and modeled as outcomes, with GrimAge AA as the predictor. Models were adjusted for age, sex, smoking status, BMI, and estimated blood cell proportions. Regression coefficients, standard errors, and *P* values were estimated for the associations between GrimAge AA and each clinical trait, and multiple testing across the 24 traits was controlled using the BH-FDR method. An additional sensitivity analysis was performed for all 24 clinical traits with further adjustment for current drinking status to account for the potential influence of alcohol consumption.

For WBC, GGT, and LDL, which showed FDR < 0.05 in the regression analyses, two-sample MR analyses were subsequently performed to evaluate potential causal relationships with GrimAge AA (Davey Smith & Hemani, 2014). Genetic associations with GrimAge AA were estimated in the main analytic cohort, whereas genetic associations with the clinical traits were estimated in the MR outcome cohort. Because the two recruitment waves comprised different participants, the exposure and outcome samples in the MR analyses did not overlap. Genome-wide association analyses were adjusted for age, sex, smoking status, BMI, and the first 10 genetic principal components. SNPs associated with GrimAge AA at *P* < 5 × 10□□ were initially selected as candidate instrumental variables, while SNPs associated with age, sex, smoking, BMI, or the corresponding clinical outcome at *P* < 5 × 10□□ were excluded. Independent instruments were subsequently selected by linkage disequilibrium clumping using r² < 0.001.

The inverse-variance weighted (IVW) method was used as the primary MR estimator. MR-Egger, weighted median, and MR-RAPS methods were additionally applied as sensitivity analyses to evaluate the robustness of the IVW estimates (Bowden et al., 2015; Zhao et al., 2020). Instrument strength was assessed using F-statistics, and I²GX was calculated to evaluate potential regression dilution in MR-Egger analyses. Heterogeneity and horizontal pleiotropy were assessed using Cochran’s Q test, the MR-Egger intercept test, and the MR-PRESSO global test (Bowden et al., 2017; Burgess & Thompson, 2011; Verbanck et al., 2018). Steiger directionality analysis was additionally performed to assess whether the genetic instruments explained greater variance in GrimAge AA than in the corresponding clinical outcome. Multiple testing across WBC, GGT, and LDL was controlled separately for each MR method using the BH-FDR procedure.

#### 2.6.5. Mediation analysis

Mediation analysis was performed to evaluate whether GrimAge AA mediated the association between environmental exposures and GGT (Fig. 1c). GGT, which showed an FDR-significant association with GrimAge AA in the observational analysis and nominal evidence of an effect in the MR analysis, was selected as the outcome, while each of the 13 environmental exposure biomarkers was evaluated separately as the exposure and GrimAge AA was specified as the mediator. All environmental exposure variables and GGT were log2-transformed prior to analysis. In the mediator model, GrimAge AA was regressed on each environmental exposure and covariates, whereas in the outcome model, GGT was regressed on the corresponding environmental exposure, GrimAge AA, and covariates. Both models were adjusted for age, sex, smoking status, BMI, and estimated blood cell proportions. Mediation effects were estimated using the R mediation package with 5,000 bootstrap simulations to estimate the average causal mediation effect (ACME), average direct effect (ADE), total effect, proportion mediated, and corresponding 95% confidence intervals (Imai et al., 2010; Tingley et al., 2014). As a sensitivity analysis, the mediation models were additionally adjusted for current drinking status to account for the potential influence of alcohol consumption on GGT.

#### 2.6.6. Mendelian randomization analysis of GrimAge AA and disease outcomes

To further investigate the potential causal effects of epigenetic aging on disease risk, two-sample MR analyses were performed using GrimAge AA as the exposure and disease-related traits as outcomes. Among the disease phenotypes available in BBJ, 38 disease outcomes were selected based on their biological relevance to aging and environmental exposure (Table S1). Genetic association estimates for GrimAge AA were obtained by meta-analyzing GWAS summary statistics from the main analytic cohort and KoGES using an inverse-variance weighted fixed-effect model to improve the precision of SNP–GrimAge AA association estimates. Effect alleles were harmonized across the two datasets prior to meta-analysis. Disease outcome summary statistics were obtained from BBJ. Although the exposure and outcome GWAS were derived from Korean and Japanese populations, respectively, previous studies have shown that these two East Asian populations are genetically closely related, with highly similar allele frequencies and LD patterns, which may reduce concerns regarding population differences in LD structure (Lim et al., 2006).

SNPs associated with GrimAge AA at *P* < 5 × 10□□ were initially selected as candidate instrumental variables. To reduce potential pleiotropic effects, SNPs associated with age, sex, smoking status, or BMI at *P* < 5 × 10□□ were excluded. SNPs associated with the corresponding disease outcome at *P* < 5 × 10□□ were also excluded. Exposure and outcome alleles were subsequently harmonized, and independent instrumental variables were selected by linkage disequilibrium clumping using r² < 0.001 with an East Asian reference panel. The IVW method was used as the primary estimator. Instrument strength and robustness were evaluated using F-statistics, Cochran’s Q test for heterogeneity, and MR-PRESSO. Multiple testing across the 38 disease outcomes was controlled using the BH-FDR method.

## 3. Results

### 3.1. Characteristics of the study population

A total of 666 participants were included in the analysis, comprising 553 individuals from environmentally vulnerable areas (exposure group) and 113 individuals from control areas (Table S2). The mean age was 70.44 years in the exposure group and 66.40 years in the control group, while the distributions of sex, BMI, and smoking status were comparable between the groups. Levels of several exposure biomarkers, including bPb, 2-HF, 1-HPH, tt-MA, MHA, and BMA, were significantly higher in the exposure group, whereas bHg was higher in the control group and the remaining biomarkers, including bCd and uCd, did not differ significantly between groups (Table S2).

We next compared epigenetic aging measures between the exposure and control groups (Fig. 2a; Table S3). GrimAge AA was significantly higher in the exposure group and remained significant after FDR correction. Hannum AA and DunedinPACE were also higher in the exposure group at nominal significance (*P* < 0.05), although these associations did not remain significant after FDR correction. epiTOC also remained significant after FDR correction; however, its level was higher in the control group than in the exposure group. No significant differences were observed for Horvath AA, Zhang AA, or PhenoAge AA.

**Fig. 2.**
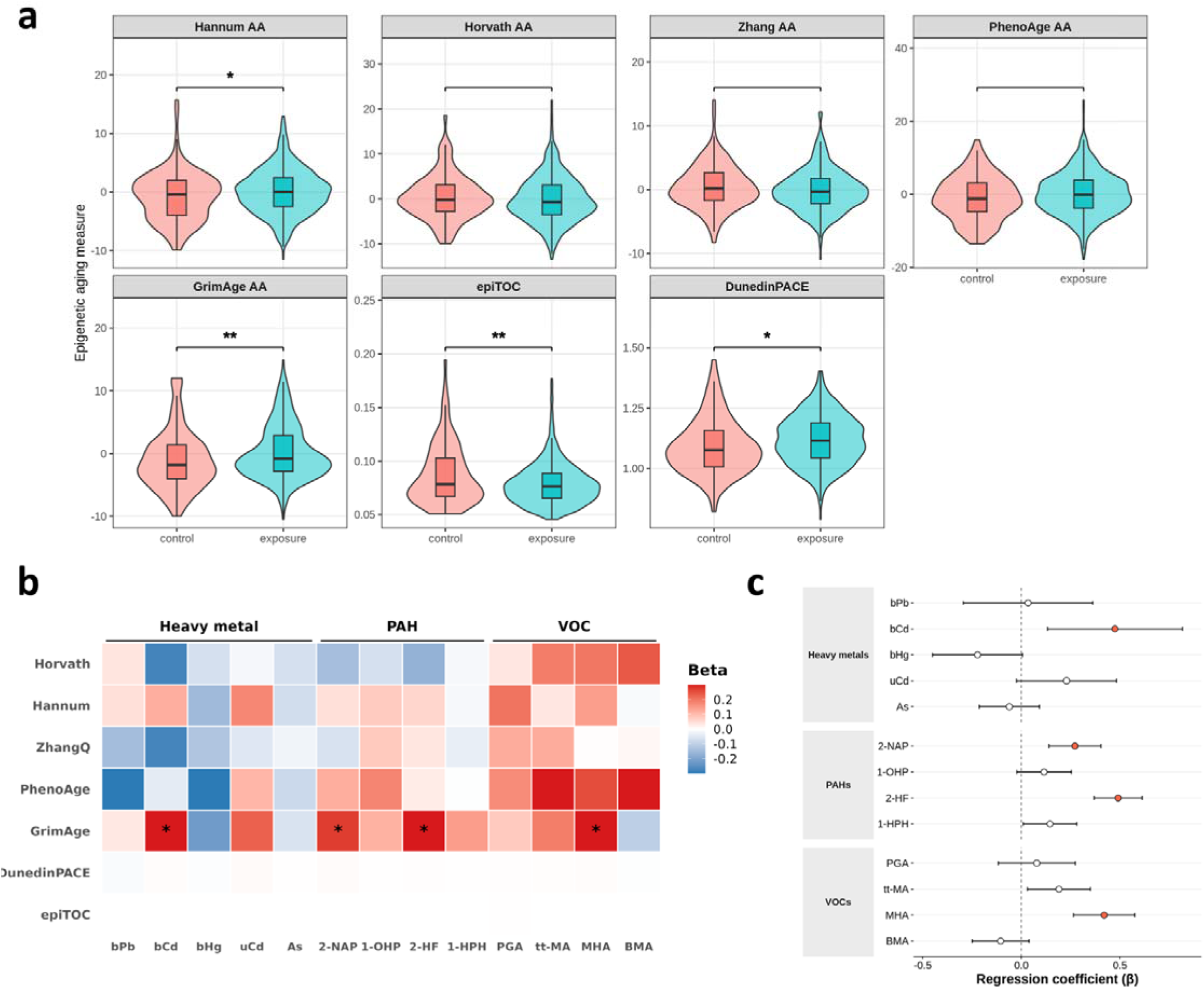
Epigenetic aging measures in the exposure and control groups and their associations with environmental exposure biomarkers. (a) Distributions of epigenetic aging measures in the exposure and control groups. Age acceleration (AA) was used for the Hannum, Horvath, Zhang, PhenoAge, and GrimAge clocks, whereas epiTOC and DunedinPACE were evaluated on their original scales. Between-group differences were assessed using Welch’s two-sample t-tests. Asterisks indicate nominal or FDR-adjusted significance (*P* < 0.05: *; FDR < 0.05: **). (b) Associations between 13 environmental exposure biomarkers and seven epigenetic aging measures. Tile colors represent regression coefficients (β) from multiple linear regression models, with red and blue indicating positive and negative associations, respectively. Asterisks indicate associations with FDR < 0.05. (c) Forest plot showing associations between individual environmental exposure biomarkers and GrimAge AA. Points represent regression coefficients (β), and error bars indicate 95% confidence intervals. Red points indicate associations with FDR < 0.05.

### 3.2. Associations between environmental exposures and epigenetic aging

Associations between the 13 environmental exposure biomarkers and seven epigenetic aging measures were evaluated using multiple linear regression (Fig. 2b, c; Fig. S3; Table S4). Significant associations after FDR correction were observed exclusively for GrimAge AA, whereas no environmental exposure biomarker was significantly associated with the other epigenetic aging measures after multiple-testing correction (Fig. S3). For GrimAge AA, blood Cd, 2-NAP, 2-HF, and MHA were positively associated with age acceleration after FDR correction (Fig. 2b, c). In particular, a two-fold increase in 2-HF was associated with a 0.491-year increase in GrimAge AA (β = 0.491 years, *P* = 1.22 × 10^−14^). The associations were robust to an alternative approach for urinary dilution adjustment using urinary concentrations without creatinine standardization and including urinary creatinine as a separate covariate, with effect estimates similar to those in the primary analysis. After FDR correction across all 13 environmental exposures, significant positive associations were observed for the urinary biomarkers 2-NAP, 2-HF, MHA, and tt-MA, as well as for blood Cd, which was unaffected by the alternative urinary dilution adjustment (Table S5). The results were also largely consistent when chronological age was modeled using a natural cubic spline, although the association with tt-MA no longer reached statistical significance after FDR correction (Table S6). The primary associations were also retained after replacing smoking status with pack-years, with 2-NAP, 2-HF, MHA, and blood Cd remaining significant after FDR correction. In analyses restricted to never-smokers, the associations of 2-HF and MHA with GrimAge AA remained significant after FDR correction, whereas the associations of 2-NAP and blood Cd were attenuated (Table S7). In sensitivity analyses stratified by sex, the positive association between 2-HF and GrimAge AA was consistently observed in both men and women, with no evidence of effect modification by sex. Several other exposure–GrimAge AA associations showed evidence of heterogeneity by sex or age; detailed exploratory results are presented in Tables S8–S11.

### 3.3. Combined effects of environmental exposures on GrimAge acceleration

To evaluate the combined effects of environmental exposures on GrimAge AA, mixture analyses were performed using qgcomp and BKMR. In the qgcomp analysis, increasing joint exposure quantiles were associated with progressively higher predicted GrimAge AA (Fig. 3a; Table S12). The overall mixture effect was positive (ψ = 0.497, 95% CI : 0.013–0.981, *P* = 0.044). Sensitivity analysis replacing urinary Cd with blood Cd yielded a consistent positive mixture effect (ψ = 0.575, 95% CI: 0.091–1.059, P = 0.020; Table S13). Positive weights were observed for several exposures, including MHA, 2-HF, 2-NAP, and 1-HPH, whereas other exposures contributed in the opposite direction (Fig. 3b; Table S14). The exposure-weight patterns were broadly similar when urinary Cd was replaced with blood Cd (Table S15).

**Fig. 3.**
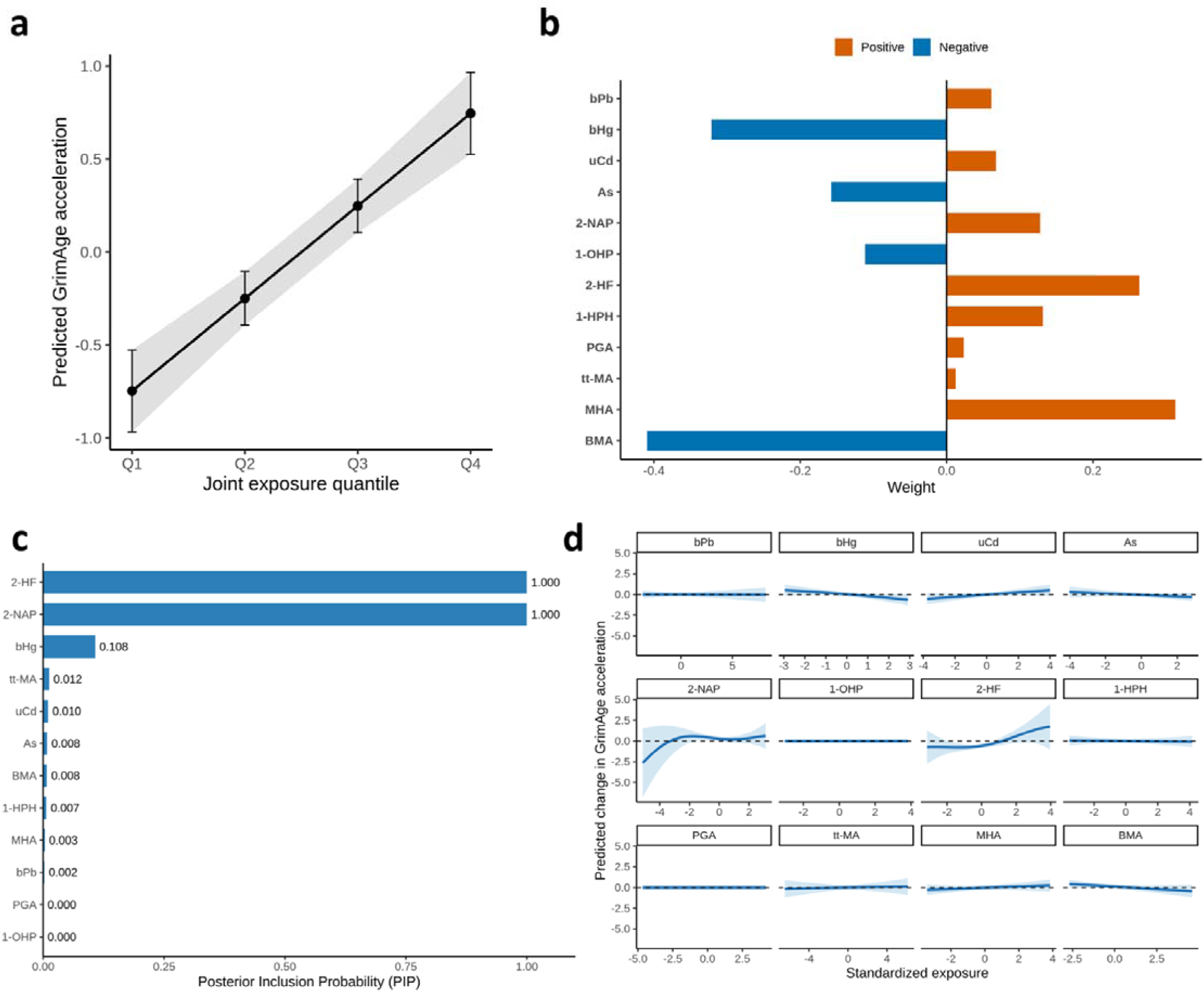
Mixture effects of environmental exposures on GrimAge acceleration. (a) Predicted GrimAge acceleration across joint exposure quantiles estimated using qgcomp. Points represent predicted values, and error bars and shaded areas indicate 95% confidence intervals. (b) Positive and negative weights of individual environmental exposures contributing to the overall mixture association in the qgcomp model. (c) PIPs of individual environmental exposures estimated using BKMR. (d) Univariate exposure–response functions for individual environmental exposures from the BKMR model, with other exposures fixed at their median levels. Solid lines represent estimated exposure–response functions, and shaded areas indicate 95% credible intervals.

BKMR analysis further identified 2-HF and 2-NAP as the predominant contributors to the mixture association, with PIPs of 1.000 for both exposures (Fig. 3c). The remaining exposure biomarkers showed substantially lower PIPs. Exposure–response functions from the BKMR model showed distinct associations of 2-HF and 2-NAP with GrimAge AA, while comparatively limited changes were observed for most other exposures (Fig. 3d). Sensitivity analysis replacing urinary Cd with blood Cd yielded consistent findings, with 2-HF and 2-NAP retaining the highest PIPs (1.000 for both) and broadly similar exposure–response patterns (Fig. S4). Together, these mixture analyses supported a combined association of environmental exposures with GrimAge AA and highlighted 2-HF and 2-NAP as important contributors to the mixture effect.

### 3.4. Associations and causal inference between GrimAge acceleration and clinical traits

Associations between GrimAge AA and 24 clinical traits were first evaluated using multiple linear regression (Fig. S5; Table S16). Among the evaluated traits, WBC and GGT showed positive associations with GrimAge AA, whereas LDL showed an inverse association, with all three associations remaining significant after FDR correction. These associations remained significant after additional adjustment for current drinking status (Table S17).

To further investigate whether the associations identified in the regression analyses were consistent with potential causal effects, two-sample MR analyses were performed for WBC, GGT, and LDL, which met the FDR < 0.05 threshold in the observational analyses (Fig. 4a; Table S18). In the primary IVW analysis, GrimAge AA showed a positive effect estimate for GGT that reached nominal significance (β = 0.067, SE = 0.030, *P* = 0.024; 7 SNPs), although the association did not remain significant after correction for the three clinical outcomes (FDR = 0.073). MR-RAPS yielded a similar positive effect estimate for GGT (β = 0.069, SE = 0.032, *P* = 0.028; FDR = 0.085), whereas the weighted median and MR-Egger estimates were not statistically significant. No evidence of an effect was observed for WBC or LDL. The mean F-statistic was 23.49, and there was no evidence of substantial heterogeneity or directional horizontal pleiotropy based on Cochran’s Q, the MR-Egger intercept test, and the MR-PRESSO global test. Steiger directionality analyses supported the hypothesized direction from GrimAge AA to each clinical outcome (all Steiger *P* < 2.1 × 10□¹□). Detailed MR estimates and instrument diagnostics are provided in Table S18, and information on the individual SNPs used as instrumental variables is provided in Tables S19–S21.

**Fig. 4.**
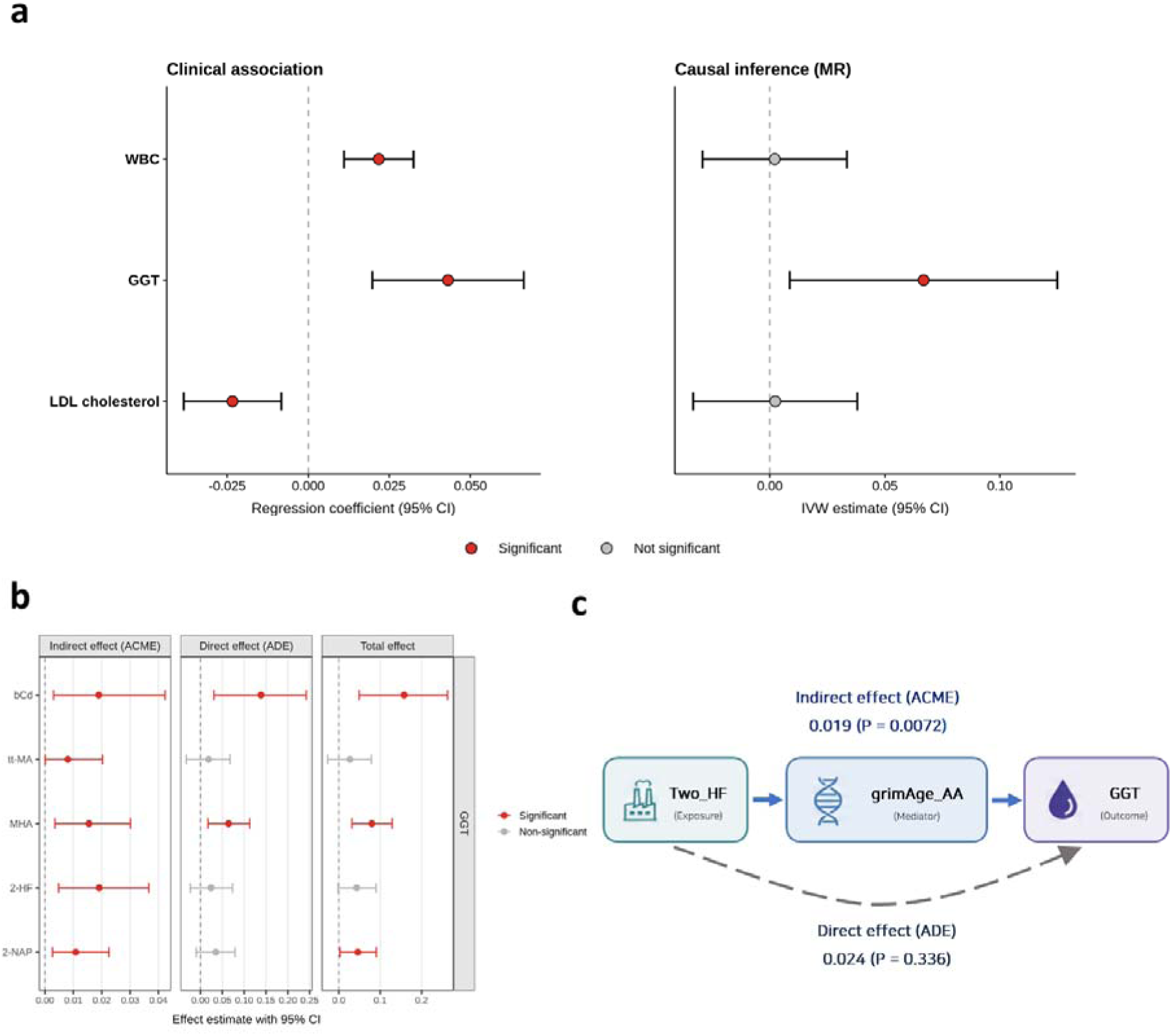
Associations of GrimAge acceleration with clinical traits and mediation of environmental exposure–GGT associations. (a) Associations of GrimAge acceleration with WBC, GGT, and LDL estimated using multiple linear regression (left) and two-sample MR using the IVW method (right). Points represent effect estimates, and error bars indicate 95% confidence intervals. Red and gray points indicate statistically significant and non-significant associations, respectively. (b) Mediation effects of environmental exposures on GGT through GrimAge acceleration. Points and error bars represent the estimated indirect effects (ACME), direct effects (ADE), and total effects with 95% confidence intervals. (c) Representative mediation pathway for the association between 2-HF and GGT through GrimAge acceleration, showing the indirect (ACME) and direct (ADE) effects.

### 3.5. Mediation of the association between environmental exposures and GGT by GrimAge acceleration

Given the significant observational association between GrimAge AA and GGT and the nominally significant IVW MR estimate, mediation analyses were subsequently performed to investigate whether GrimAge AA mediated the associations between environmental exposures and GGT. Each of the 13 environmental exposure biomarkers was evaluated as an exposure, with GrimAge AA as the mediator and GGT as the outcome (Fig. S6; Table S22). Significant indirect effects were observed for five environmental exposures (2-HF, 2-NAP, bCd, MHA, and tt-MA) (Fig. 4b). Among these exposures, all except tt-MA remained significant after BH-FDR correction. These findings were robust to additional adjustment for current drinking status, with the same indirect effects remaining significant after BH-FDR correction (Table S23).

Among these associations, 2-HF is presented as a representative example in Fig. 4c. The association between 2-HF exposure and GGT was significantly mediated through GrimAge AA (ACME = 0.019, *P* = 0.0072), whereas the direct effect was not statistically significant (ADE = 0.024, *P* = 0.336).

### 3.6. Potential causal effects of GrimAge acceleration on disease outcomes

MR analyses were performed to investigate the potential causal effects of GrimAge AA on a broad range of disease outcomes. Among the 38 disease outcomes evaluated, four showed nominally significant associations (*P* < 0.05): diabetic nephropathy (β = 0.1812, *P* = 0.0025), osteoporosis (β = 0.0282, *P* = 0.0054), cardiac valvular disease (β = −0.0545, *P* = 0.0074), and dilated cardiomyopathy (β = 0.1213, *P* = 0.0388). After FDR correction, diabetic nephropathy, osteoporosis, and cardiac valvular disease showed suggestive associations (FDR = 0.0933), although none of the disease associations met the conventional FDR < 0.05 threshold. The associations were positive for diabetic nephropathy and osteoporosis and inverse for cardiac valvular disease. Detailed MR results for all evaluated disease outcomes, including instrument strength, heterogeneity, and MR-PRESSO statistics, are provided in Tables S24–S25. Detailed information on the SNPs selected as instrumental variables, including their associations with the exposure and outcome traits, is presented in Tables S26–S29.

## 4. Discussion

In this community-based study of 666 older adults, including 553 residents of environmentally vulnerable areas and 113 controls, environmental exposure was associated with accelerated epigenetic aging and downstream physiological alterations. Four findings were central. First, among seven epigenetic aging measures, GrimAge AA showed the most consistent signal across both the exposed-versus-control comparison and biomarker-based analyses. Second, PAH biomarkers, particularly 2-HF and 2-NAP, emerged as reproducible correlates of GrimAge AA across individual- and mixture-exposure models. Third, GrimAge AA was associated with WBC, GGT, and LDL, but MR provided nominal support for a directional effect only for GGT; mediation analyses further identified indirect associations of 2-HF, 2-NAP, bCd, and MHA with GGT through GrimAge AA that remained significant after FDR correction. Finally, exploratory disease MR suggested potential associations with diabetic nephropathy, osteoporosis, and cardiac valvular disease at FDR < 0.10. Together, these findings suggest that pollutant-associated GrimAge AA may reflect physiologically relevant biological alterations.

The specificity of the exposure signal to GrimAge is biologically informative. In the regional comparison, Hannum AA and DunedinPACE were nominally significant, whereas epiTOC remained significant after FDR correction but was higher in controls. In quantitative exposure models, only GrimAge AA retained associations after FDR correction, specifically with bCd, 2-NAP, 2-HF, and MHA. First-generation clocks such as Horvath and Hannum were optimized primarily to predict chronological age, whereas GrimAge was trained to predict mortality using DNAm surrogates of smoking pack-years and mortality-related plasma proteins (Hannum et al., 2013; Horvath, 2013; Lu et al., 2019). These components capture smoking-related, inflammatory, vascular, renal, and metabolic information, potentially explaining the greater sensitivity of GrimAge to combustion- and toxicant-related exposures. The opposite-direction epiTOC finding further suggests that mitotic age and mortality-related biological age represent distinct aging domains.

This interpretation is consistent with previous environmental epigenetic-aging studies. Occupational and community PAH exposure has been associated with accelerated DNAm aging (Li et al., 2018), and PAH-related exposure from household smoky-coal combustion was particularly associated with GrimAge AA among never-smoking women (Blechter et al., 2023). Occupational benzene and trichloroethylene exposure has also been related to epigenetic aging, although affected clocks differed across exposures and populations (Van der Laan et al., 2022). For metals, cadmium and a nonessential-metal mixture containing cadmium were associated with greater GrimAge AA and faster DunedinPACE in American Indian populations (Boyer et al., 2023), while an exposome-wide analysis of 2,346 US adults identified serum cadmium as most strongly associated with GrimAge AA (Khodasevich et al., 2025). Together, these studies support exposure-specific heterogeneity across epigenetic clocks and suggest that GrimAge may be particularly responsive to environmental exposures involving smoking-related, inflammatory, and multisystem stress.

The PAH findings were particularly consistent across analytical approaches. In single-pollutant models, 2-HF showed a strong association with GrimAge AA (β = 0.491, *P* = 1.22 × 10^−14^), which was observed in both men and women without evidence of sex interaction. Qgcomp estimated a positive joint mixture effect, with positive weights for MHA, 2-HF, 2-NAP, and 1-HPH, while BKMR assigned posterior inclusion probabilities of 1.000 to both 2-HF and 2-NAP. Their convergence across conventional regression, qgcomp, and BKMR therefore identifies 2-HF and 2-NAP as prominent contributors to the observed exposure signal. Because these urinary metabolites reflect exposure to fluorene, naphthalene, and correlated combustion sources, however, the findings are more appropriately interpreted as markers of PAH-related exposure rather than evidence of toxicity from the measured metabolites themselves.

Several mechanisms may connect PAH exposure with GrimAge AA. Parent PAHs can activate the aryl hydrocarbon receptor and induce *CYP1A1* and *CYP1B1*, while metabolic activation generates electrophilic intermediates and o-quinones capable of forming DNA adducts and generating reactive oxygen species, DNA-strand breaks, and oxidative DNA damage (Endo et al., 2008; Park et al., 2005; Park et al., 2009; Xue & Warshawsky, 2005). Consistent with these mechanisms, urinary hydroxylated PAHs have been associated with oxidative nucleic-acid damage, lipid peroxidation, C-reactive protein, and WBC count in human studies (Alshaarawy et al., 2013; Kim et al., 2003; Lu et al., 2016; Zhang et al., 2020). Notably, 2/3-hydroxyfluorene and 2-hydroxynaphthalene were recently identified as important contributors to urinary oxidative nucleic-acid damage biomarkers (Cao et al., 2024), while experimental chronic PAH exposure has produced persistent oxidative DNA damage and genomic instability (Bai et al., 2017).

Recurrent oxidative and inflammatory stress may subsequently alter DNA methylation. Oxidative DNA damage can recruit DNMT1- and DNMT3B-containing repressive complexes to damaged chromatin (O’Hagan et al., 2011), while human PAH exposure has been associated with methylation changes in *OGG1* and *AHRR*, age-related loci including *FHL2* and *ELOVL2*, and accelerated DNAm aging (Chen et al., 2024; Fu et al., 2019; Li et al., 2018). This mechanism is particularly compatible with GrimAge, which incorporates DNAmPACKYRS and DNAm surrogates for proteins involved in inflammatory, vascular, renal, stress-response, and tissue-remodeling processes (Lu et al., 2019). Consistently, smoky-coal combustion exposure has been associated with GrimAge AA even among never-smoking women (Blechter et al., 2023).

The accompanying bCd and MHA findings, together with the more tentative tt-MA signal, broaden the interpretation from PAHs alone to a multi-pollutant aging signal. Cadmium is retained in the body, promotes mitochondrial dysfunction, glutathione depletion, inflammation, and oxidative injury, and has now been linked to GrimAge AA in several populations (Boyer et al., 2023; Khodasevich et al., 2025). tt-MA and MHA are urinary biomarkers used to index benzene and xylene exposure, respectively, and therefore may represent additional components of an inhaled or occupationally patterned mixture rather than independent causal agents. Residential VOC biomarker studies have associated low-level exposure with hepatocyte injury and altered liver enzymes, while occupational benzene exposure has been associated with epigenetic age acceleration (Van der Laan et al., 2022; Wahlang et al., 2022). Shared mechanisms—including reactive metabolite formation, glutathione consumption, oxidative stress, and inflammatory signaling—could therefore explain why chemically distinct biomarkers converged on GrimAge AA and GGT. At the same time, the short half-lives and source nonspecificity of several urinary metabolites require caution when assigning effects to individual parent compounds.

The clinical-trait analyses support the biological and clinical relevance of GrimAge AA beyond its role as a methylation-derived computational score. GrimAge AA was positively associated with WBC and GGT and inversely associated with LDL after FDR correction. The WBC association is compatible with systemic inflammatory or immune-cell activation, one of the pathways through which PAHs and metals may affect aging. By contrast, the inverse LDL association was not supported by MR and may reflect lipid-lowering treatment, comorbidity, nutrition, frailty, or survival-related selection in this older population; it should not be interpreted as a protective effect of accelerated aging. Similarly, the observational association with WBC was not supported by MR. GGT was therefore the most coherent downstream phenotype because it was supported by both the observational association and the MR analysis.

GGT provides a biologically plausible link between pollutant exposure and GrimAge-associated physiological dysregulation. Beyond its conventional role as a hepatobiliary marker, GGT participates in extracellular glutathione metabolism and may increase in response to oxidative stress (Lee et al., 2004; Zhang & Forman, 2009). This is particularly relevant to the PAH findings: urinary 2-HF has been associated with elevated GGT in US adults (Mallah et al., 2023), while a PAH mixture was positively associated with GGT in Korean adults, with 2-NAP and 2-HF among the largest contributors (Ryu & Hong, 2024). Extending these findings, our study links internal PAH exposure in residents of environmentally vulnerable areas to GGT through GrimAge AA, suggesting that exposure-related epigenetic aging may represent an intermediate biological process connecting pollutant exposure with physiological dysregulation. Inflammation and oxidative stress have also been implicated in PAH–liver-function associations, with 2-HF contributing prominently to elevated GGT (Zhou et al., 2024), and these metabolites have been associated with ALT elevation or fatty-liver-related phenotypes (Choi et al., 2023; Lee et al., 2025). Cadmium has likewise been associated with higher GGT and other liver-injury markers (Kim et al., 2021; Yan et al., 2022). Moreover, higher GGT has prospectively predicted type 2 diabetes, chronic kidney disease, and osteoporotic fracture in Korean populations (Kim et al., 2014; Lee et al., 2020; Ryu et al., 2007), suggesting that it may reflect broader redox and metabolic dysregulation rather than liver injury alone.

The mediation results provide the most direct synthesis of these findings. Significant indirect effects were observed for 2-HF, 2-NAP, bCd, and MHA in relation to GGT through GrimAge AA. For 2-HF, the estimated average causal mediation effect was 0.019 (*P* = 0.0072), whereas the average direct effect was 0.024 (*P* = 0.336). This pattern is compatible with a pathway in which repeated toxicant-related oxidative and inflammatory stress becomes embedded in a mortality-related DNAm signature, which then tracks or contributes to a glutathione-dependent hepatometabolic response reflected by GGT. The result is strongest for 2-HF and 2-NAP because both were supported in the individual and mixture analyses, and because independent studies have linked them to GGT or related liver outcomes. Importantly, a nonsignificant direct effect is not proof of complete mediation, and GrimAge should not be treated as a single molecular effector. It is more plausibly an integrative record of smoking-like exposure, inflammation, protein regulation, and cumulative organ stress. Moreover, the MR result supports the direction from GrimAge AA to GGT but does not validate the exposure-to-GrimAge portion of the mediation model. Accordingly, the pathway should be interpreted as biologically coherent and hypothesis-generating rather than as definitive temporal causation.

Exploratory disease MR provided additional, although suggestive, evidence of clinical relevance. Among 38 outcomes, diabetic nephropathy (β = 0.1812, *P* = 0.0025), osteoporosis (β = 0.0282, *P* = 0.0054), and cardiac valvular disease (β = −0.0545, *P* = 0.0074) showed suggestive associations with GrimAge AA at FDR < 0.10, although none met FDR < 0.05. Whereas the associations with diabetic nephropathy and osteoporosis were positive, the association with cardiac valvular disease was inverse; given the limited evidence supporting a protective relationship between accelerated epigenetic aging and valvular disease, this finding should be interpreted cautiously and requires independent replication. Biological aging and diabetic kidney disease share mitochondrial dysfunction, oxidative stress, inflammation, and cellular senescence; human and experimental studies have demonstrated increased senescence and impaired mitochondrial quality control in diabetic nephropathy (Chen et al., 2018; Kitada et al., 2014; Verzola et al., 2008). DNAm aging measures have also been associated with renal dysfunction and progression of diabetic complications (Matías-García et al., 2021; Vetter et al., 2023). Similarly, cellular senescence contributes to age-related bone loss (Farr et al., 2017), and a recent twin study identified GrimAge AA as a consistent epigenetic-aging correlate of osteoporosis and fracture phenotypes (Soerensen et al., 2025). A previous bidirectional MR study found no evidence for causal effects of GrimAge AA on bone mineral density at the femoral neck, forearm, or lumbar spine; however, the reverse analysis suggested that lower lumbar spine bone mineral density may contribute to accelerated GrimAge (Liang et al., 2023). Thus, these disease findings require replication and should be interpreted as suggestive links between accelerated epigenetic aging and age-related disease rather than definitive causal relationships.

This study has several notable strengths. First, it evaluated seven epigenetic aging measures that capture distinct dimensions of biological aging, allowing the relative specificity of the GrimAge signal to be assessed rather than assumed. Second, the exposure findings were examined at complementary levels: regional exposure-group comparisons, individual biomarker associations, and multi-pollutant mixture models. The concordance of results across multivariable regression, quantile g-computation, and BKMR—particularly for 2-HF and 2-NAP—strengthens confidence that these findings were not driven by a single modeling approach. Third, the integration of clinical-trait analyses, mediation analyses, and Mendelian randomization extended the investigation beyond exposure–DNAm associations to evaluate their potential physiological and disease relevance. Finally, the focus on older Korean adults residing in environmentally vulnerable areas addresses a population in which cumulative exposure burdens and susceptibility to aging-related effects may be especially relevant.

Several limitations should be considered. First, exposure biomarkers, DNAm, and clinical measurements were largely assessed cross-sectionally, limiting temporal and causal interpretation, while single urine measurements may not adequately represent usual exposure to rapidly cleared PAHs and VOCs. Second, residual confounding by smoking, second-hand smoke, alcohol use, diet, medication, occupation, socioeconomic conditions, and pre-existing disease remains possible; this is particularly relevant because GrimAge incorporates a DNAm smoking surrogate. Third, the exposed group was older than controls, and residual age-related differences may remain despite the use of age-acceleration measures and covariate adjustment. In addition, the relatively small control group (n = 113) limited the statistical power of the regional comparisons. Fourth, although multiple epigenetic aging measures were evaluated, more recently developed measures, such as GrimAge2 and PCGrimAge, were not included, which may limit the comprehensiveness of our assessment of exposure-related epigenetic aging. Fifth, mediation analysis relies on strong no-unmeasured-confounding assumptions that cannot be verified in this observational setting. Sixth, DNA methylation was measured using two EPIC array versions, and although batch effects were corrected, residual array-related variation cannot be fully ruled out. Finally, although we conducted sensitivity analyses to assess instrument strength, heterogeneity, and potential horizontal pleiotropy, the genetic instruments for GrimAge AA were derived from relatively small GWAS (n = 666 for the clinical-trait MR and n = 1,892 for the disease MR) using a relaxed significance threshold (*P* < 5 × 10□□). Therefore, winner’s curse, weak-instrument bias, and residual horizontal pleiotropy cannot be excluded. In the clinical-trait MR analyses, the nominal association observed for GGT did not remain significant after correction for multiple testing across the three clinical outcomes. Similarly, the disease MR findings achieved only suggestive significance at FDR < 0.10. Differences in ancestry, phenotype definitions, and genetic architecture may further limit transportability, underscoring the need for independent replication.

## 5. Conclusions

This study indicates that environmental pollutant exposure—most consistently PAH exposure indexed by 2-HF and 2-NAP—is associated with accelerated mortality-related epigenetic aging. GrimAge was the only aging measure that showed a coherent adverse-exposure signal across regional and biomarker-based analyses, and its associations with GGT, mediation of several pollutant–GGT relationships, and suggestive MR links to diabetic nephropathy and osteoporosis support biological and clinical relevance. These results support a broader framework linking environmental pollutants to accelerated epigenetic aging and, in turn, to adverse physiological change, with GGT serving as one downstream redox–hepatometabolic indicator rather than a terminal endpoint. Longitudinal studies with repeated exposure measurements, serial DNAm profiling, component-level GrimAge analyses, and independently replicated genetic instruments are needed to determine whether reducing PAH and co-pollutant exposure can slow biological aging and lessen aging-related disease risk.

## CRediT authorship contribution statement

**Hyeongyu Shin:** Conceptualization, Methodology, Formal analysis, Data curation, Visualization, Writing – original draft, Writing – review & editing. **Jeeyoung Kim:** Investigation. **Euikyoung Song:** Data curation. **Nam-Eun Kim:** Data curation. **Min Heo:** Data curation. **Youngwoong Kim:** Data curation. **Jeongmin Oh:** Data curation. **Yong Min Cho:** Investigation, Funding acquisition. **Young-Seoub Hong:** Investigation, Project administration, Funding acquisition. **Woo Jin Kim:** Writing – review & editing, Investigation, Project administration, Funding acquisition. **Sungho Won:** Supervision, Writing – review & editing, Funding acquisition.

## Acknowledgements

This work was supported by a grant from the Institute of Information & communications Technology Planning & Evaluation (IITP), funded by the Korean government (MSIT) (No. RS-2025-25441324, Development of AI-Enabled, Phenome Data-Driven Precision Health Prediction and Management Solutions for Customized Life Coaching of Healthy Adults), and a National Research Foundation (NRF) grant (NRF-2021R1A5A1033157) funded by the Korean government (MSIT). This research was supported by a grant from the Korea Health Technology R&D Project through the Korea Health Industry Development Institute (KHIDI), funded by the Ministry of Health & Welfare, Republic of Korea (grant number: RS-2025-24535069).

This study used biomedical and research resources, including genetic and health information, provided by CODA (Clinical & Omics Data Archive), the Agency for Disease Control and Prevention, Republic of Korea (CODA_S2500024-01). Statistical analyses were supported by the national supercomputing center with supercomputing resources and technical support (KSC-2024-CRE-0514 and KSC-2025-CRE-0271). Large data transfer was supported by KREONET, which is managed and operated by KISTI.

## Data Availability

Raw data generated from the FROM study are available from the corresponding author (WJK) upon reasonable request. Additional data used in this study were sourced from the Korean Genome and Epidemiology Study (KoGES), conducted by the National Institute of Health, Korea Disease Control and Prevention Agency. Summary-level results supporting the findings of this study are available in the main text or the supplementary materials.

