## Supplementary Materials for "Environmental Chemical Exposures and Accelerated Epigenetic Aging: Links to Metabolic and Age-Related Health Outcomes"

Hyeongyu Shin *et al.*

*Sungho Won.

*Woo Jin Kim.

**This PDF file includes:**

Figs. S1 to S6

Tables S1 to S29

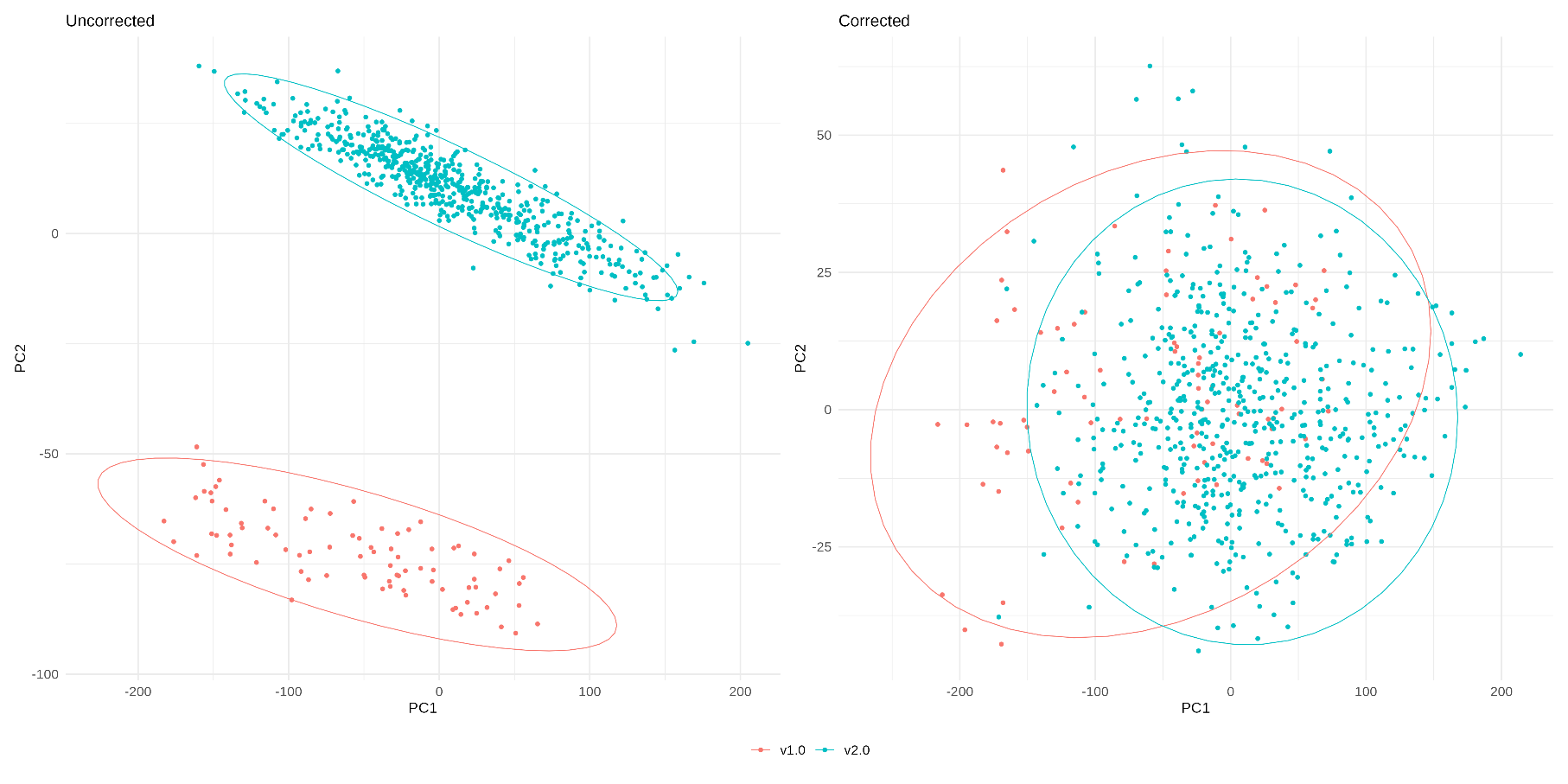
**Supplementary Figure S1. Assessment of array-version effects before and after ComBat.** Plots show the first two components of DNA methylation data before (Uncorrected) and after (Corrected) ComBat correction. Samples are colored according to EPIC array version (v1.0 and v2.0), and ellipses indicate the distribution of samples within each array version. Clear separation between EPIC v1.0 and v2.0 samples before correction was substantially reduced after ComBat correction.

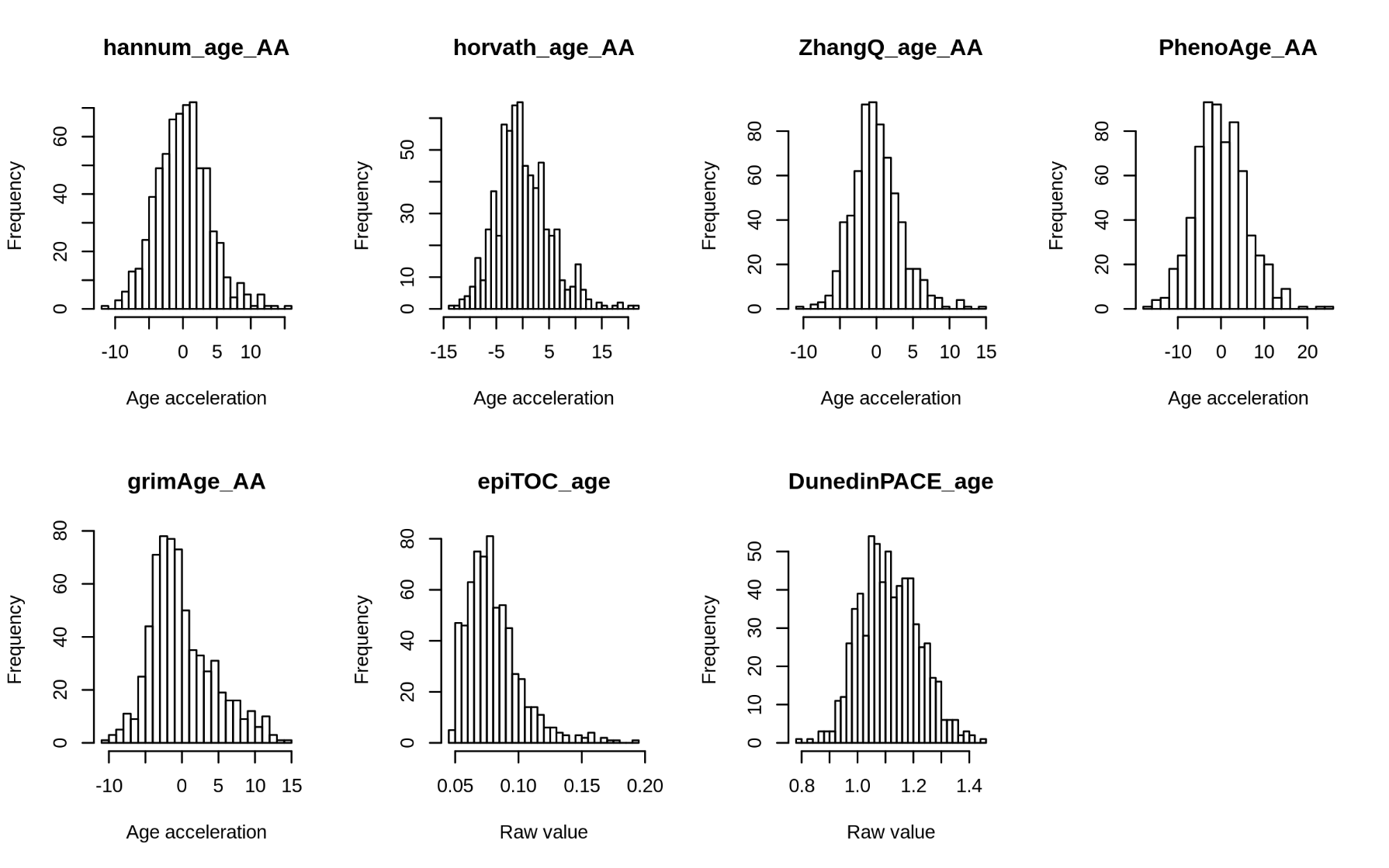
**Supplementary Figure S2. Distributions of epigenetic aging measures in the study population.** Histograms show the distributions of Hannum age acceleration (AA), Horvath AA, Zhang AA, PhenoAge AA, GrimAge AA, epiTOC, and DunedinPACE among the 666 study participants. AA measures represent residuals from linear regression of epigenetic age on chronological age; epiTOC and DunedinPACE are presented as raw values.

**Supplementary Figure S3. Associations between environmental exposure biomarkers and epigenetic aging measures other than GrimAge age acceleration.** Forest plots show regression coefficients (β) and 95% confidence intervals from multivariable linear regression models for associations of heavy metals, polycyclic aromatic hydrocarbon (PAH) metabolites, and volatile organic compound (VOC) metabolites with Horvath AA, Hannum AA, Zhang AA, PhenoAge AA, epiTOC, and DunedinPACE. To facilitate visualization across epigenetic aging measures with different scales, regression coefficients and their standard errors were rescaled to the standard deviation of each outcome. Exposure biomarkers were log2-transformed, and models were adjusted for age, sex, smoking status, BMI, and estimated blood cell proportions. The dashed vertical line indicates the null value.

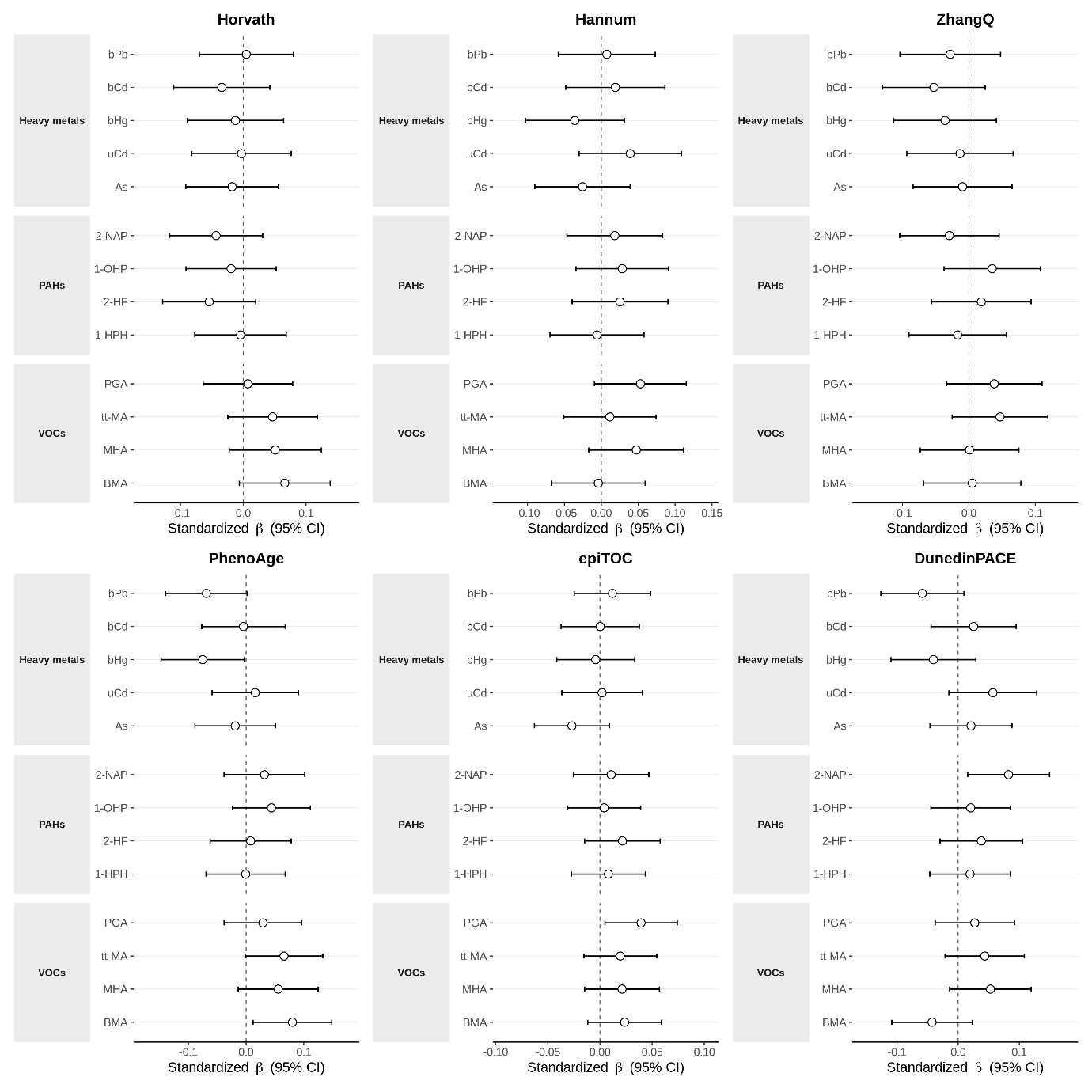

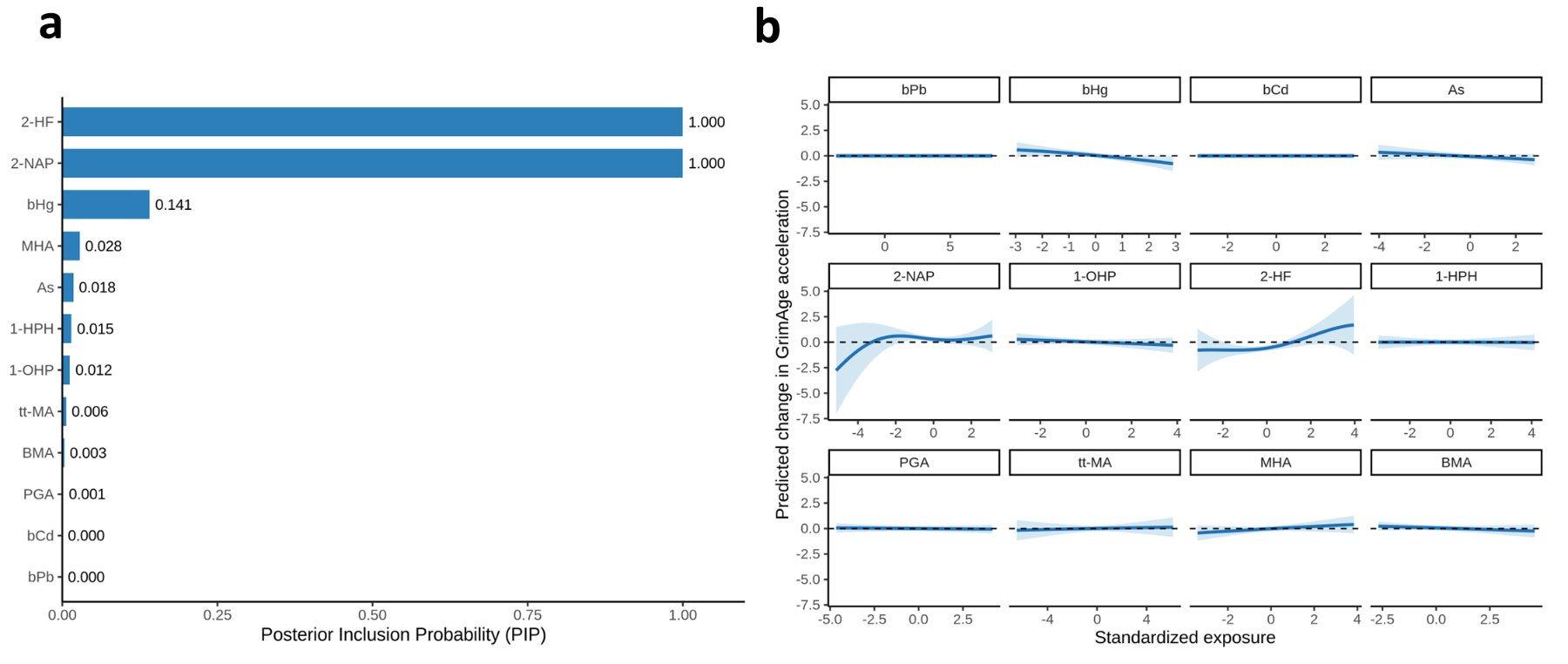
**Supplementary Fig. S4. Sensitivity analysis of Bayesian kernel machine regression (BKMR) replacing urinary cadmium with blood cadmium.** (a) PIPs of individual environmental exposures estimated using BKMR. (b) Univariate exposure–response functions for individual environmental exposures from the BKMR model, with other exposures fixed at their median levels. Solid lines represent estimated exposure–response functions, and shaded areas indicate 95% credible intervals.

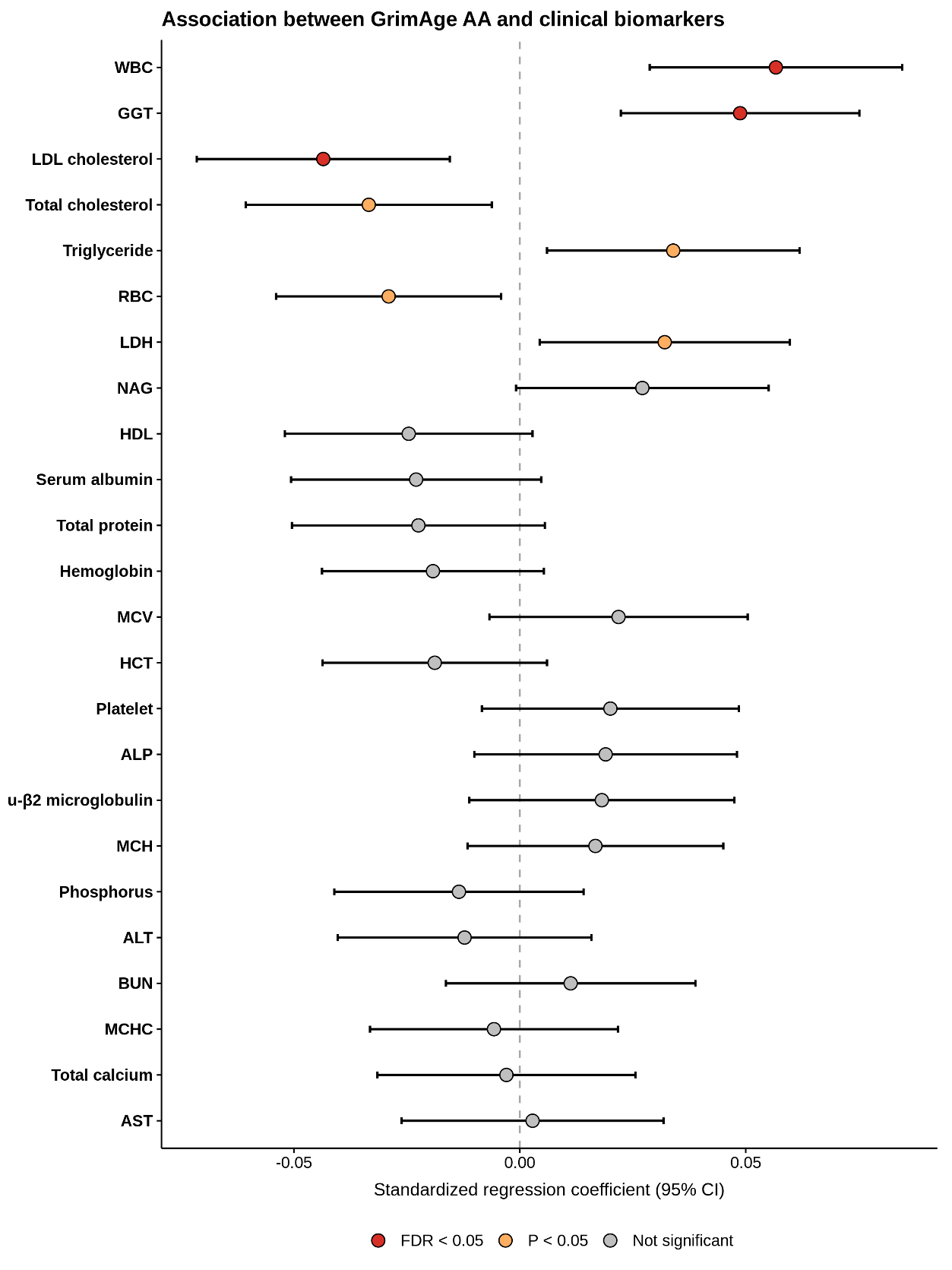
**Supplementary Figure S5. Associations between GrimAge age acceleration and clinical traits.** Forest plots show regression coefficients (β) and 95% confidence intervals from multivariable linear regression models, with each log2-transformed clinical trait as the outcome and GrimAge AA as the predictor. To facilitate visualization across clinical traits with different scales, regression coefficients and their standard errors were rescaled to the standard deviation of each outcome. Models were adjusted for age, sex, smoking status, BMI, and estimated blood cell proportions. Red, orange, and gray points indicate associations with FDR < 0.05, nominal *P* < 0.05, and non-significant associations, respectively.

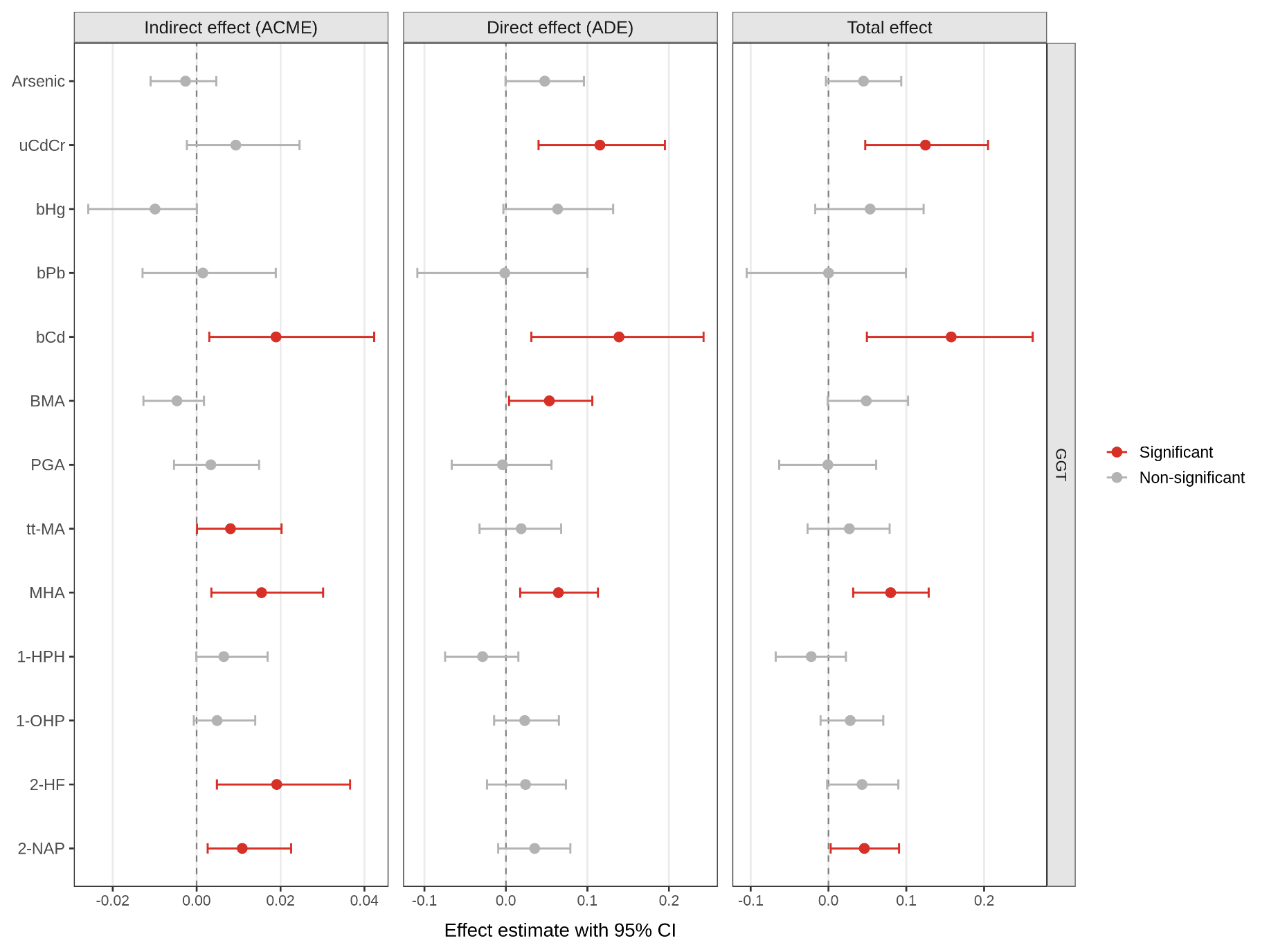
**Supplementary Figure S6. Mediation of associations between environmental exposure biomarkers and GGT through GrimAge age acceleration.** Forest plots show the average causal mediation effect (ACME), average direct effect (ADE), and total effect for each environmental exposure biomarker, with GrimAge AA specified as the mediator and log2-transformed GGT as the outcome. Estimates and 95% confidence intervals were obtained from mediation analyses adjusted for age, sex, smoking status, BMI, and estimated blood cell proportions using 5,000 bootstrap simulations. Red and gray points indicate statistically significant and non-significant effects, respectively. The dashed vertical line indicates the null value.

**Supplementary Table S1. Disease outcomes included in the Mendelian randomization analysis.** Disease outcomes were selected from the BioBank Japan (BBJ) resource based on their biological relevance to aging and environmental exposure and were grouped by disease category. **Supplementary Table S2. Descriptive characteristics of the study participants.** Values are presented as mean (standard deviation [SD]) for age, BMI, and epigenetic age measures; median (interquartile range [IQR]) for blood and urinary exposure biomarkers; and number (%) for categorical variables. Urinary biomarkers were creatinine-adjusted and are expressed as μg/g creatinine. Differences between the exposure and control groups were assessed using the Wilcoxon rank-sum test for continuous variables and the chi-square test for categorical variables.

**Supplementary Table S3. Comparison of epigenetic aging measures between the exposure and control groups.** Group-specific values are presented as mean and SD. Differences were assessed using Welch’s two-sample t-test. False discovery rate (FDR)-adjusted P values were calculated using the Benjamini–Hochberg procedure.

**Supplementary Table S4. Associations between environmental exposure biomarkers and epigenetic aging measures.** Results were estimated using separate multivariable linear regression models for each exposure biomarker and epigenetic aging measure, adjusted for age, sex, smoking status, BMI, and estimated blood cell proportions. Exposure biomarkers were log2-transformed before analysis. FDR-adjusted P values were calculated across the 13 exposure biomarkers separately for each epigenetic aging measure.

**Supplementary Table S5. Sensitivity analysis of associations between environmental exposure biomarkers and GrimAge age acceleration using an alternative urinary dilution adjustment.** Separate multivariable linear regression models were fitted for each exposure biomarker and were adjusted for age, sex, smoking status, BMI, and estimated blood cell proportions. Exposure biomarkers were log2-transformed before analysis. Urinary exposure biomarkers were analyzed using concentrations without creatinine standardization, with urinary creatinine included as a separate covariate, whereas blood biomarkers were analyzed as in the primary analysis. FDR-adjusted P values were calculated across all 13 exposure biomarkers.

**Supplementary Table S6. Sensitivity analysis of associations between environmental exposure biomarkers and GrimAge age acceleration using nonlinear adjustment for chronological age.** Separate multivariable linear regression models were fitted for each exposure biomarker, with chronological age modeled using a natural cubic spline with 3 degrees of freedom instead of a linear age term. Models were additionally adjusted for sex, smoking status, BMI, and estimated blood cell proportions. Exposure biomarkers were log2-transformed before analysis. FDR-adjusted P values were calculated across all 13 exposure biomarkers.

**Supplementary Table S7. Sensitivity analyses of associations between environmental exposure biomarkers and GrimAge age acceleration according to smoking history.** Separate multivariable linear regression models were fitted for each exposure biomarker. In the pack-years analysis, smoking status in the primary model was replaced with cumulative smoking exposure quantified as pack-years, with adjustment for age, sex, BMI, and estimated blood cell proportions. One participant with missing information required to calculate pack-years was excluded, resulting in 665 participants. In the never-smoker analysis, the models were restricted to participants who reported never smoking (n = 471) and were adjusted for age, sex, BMI, and estimated blood cell proportions. Exposure biomarkers were log2-transformed before analysis. FDR-adjusted P values were calculated across all 13 exposure biomarkers separately for each sensitivity analysis.

**Supplementary Table S8. Sex-stratified associations between environmental exposure biomarkers and GrimAge age acceleration.** Separate multivariable linear regression models were fitted for men and women and were adjusted for age, smoking status, BMI, and estimated blood cell proportions. Exposure biomarkers were log2-transformed before analysis. FDR-adjusted P values were calculated across the exposure biomarkers within each sex stratum.

**Supplementary Table S9. Sex interaction analyses for associations between environmental exposure biomarkers and GrimAge age acceleration.** Interaction terms between each log2-transformed exposure biomarker and sex were evaluated in multivariable linear regression models adjusted for age, sex, smoking status, BMI, and estimated blood cell proportions. Men were used as the reference group. FDR-adjusted P values were calculated across the exposure biomarkers.

**Supplementary Table S10. Age-stratified associations between environmental exposure biomarkers and GrimAge age acceleration.** Separate multivariable linear regression models were fitted for participants aged <70 and ≥70 years and were adjusted for age, sex, smoking status, BMI, and estimated blood cell proportions. Exposure biomarkers were log2-transformed before analysis. FDR-adjusted P values were calculated across the exposure biomarkers within each age stratum.

**Supplementary Table S11. Age interaction analyses for associations between environmental exposure biomarkers and GrimAge age acceleration.** Interaction terms between each log2-transformed exposure biomarker and continuous age were evaluated in multivariable linear regression models adjusted for age, sex, smoking status, BMI, and estimated blood cell proportions. FDR-adjusted P values were calculated across the exposure biomarkers.

**Supplementary Table S12. Overall association of the environmental exposure mixture with GrimAge age acceleration estimated using quantile g-computation.** The estimate represents the change in GrimAge age acceleration associated with a simultaneous one-quartile increase in all mixture components. The model was adjusted for age, sex, smoking status, BMI, and estimated blood cell proportions. Confidence intervals were estimated using 1,000 bootstrap iterations.

**Supplementary Table S13. Sensitivity analysis of the overall association between the environmental exposure mixture and GrimAge age acceleration estimated using quantile g-computation.** In this analysis, urinary cadmium (Cd) was replaced with blood Cd, while all other exposure variables and covariates were retained. The estimate represents the change in GrimAge age acceleration associated with a simultaneous one-quartile increase in all mixture components. The model was adjusted for age, sex, smoking status, BMI, and estimated blood cell proportions. Confidence intervals were estimated using 1,000 bootstrap iterations.

**Supplementary Table S14. Directional weights of environmental exposure biomarkers in the quantile g-computation analysis.** Positive and negative weights represent the relative contributions of individual mixture components to the overall positive and negative associations with GrimAge age acceleration, respectively.

**Supplementary Table S15. Sensitivity analysis of directional weights of environmental exposure biomarkers in the quantile g-computation analysis.** In this analysis, urinary cadmium (Cd) was replaced with blood Cd, while all other exposure variables and covariates were retained. Positive and negative weights represent the relative contributions of individual mixture components to the overall positive and negative associations with GrimAge age acceleration, respectively.

**Supplementary Table S16. Associations between GrimAge age acceleration and clinical traits.** Results were estimated using multivariable linear regression models with GrimAge age acceleration as the predictor and each log2-transformed clinical trait as the outcome. Models were adjusted for age, sex, smoking status, BMI, and estimated blood cell proportions. FDR-adjusted P values were calculated across the evaluated clinical traits.

**Supplementary Table S17. Sensitivity analysis of the associations between GrimAge age acceleration and clinical traits with additional adjustment for current drinking status.** Results were estimated using multivariable linear regression models with GrimAge age acceleration as the predictor and each log2-transformed clinical trait as the outcome. Models were adjusted for age, sex, smoking status, BMI, estimated blood cell proportions, and current drinking status. FDR-adjusted P values were calculated across the evaluated clinical traits.

**Supplementary Table S18. Two-sample Mendelian randomization estimates for the effects of GrimAge age acceleration on GGT, LDL, and WBC.** The inverse-variance weighted (IVW) method was used as the primary estimator, with MR-Egger, weighted median, and MR-RAPS analyses performed as sensitivity analyses. Instrument strength and robustness were assessed using the mean F-statistic, I²GX statistic, Cochran’s Q test, MR-PRESSO global test, MR-Egger intercept test, and Steiger directionality analysis. False discovery rate (FDR)-adjusted P values were calculated across the three clinical outcomes separately for each MR method using the Benjamini–Hochberg procedure. I²GX was used to assess potential regression dilution in MR-Egger analyses due to measurement error in SNP–exposure associations. In the Steiger analysis, the reported direction indicates whether the genetic instruments explained greater variance in GrimAge age acceleration than in the corresponding clinical outcome, and the Steiger P value represents the statistical evidence supporting the inferred direction.

**Supplementary Table S19. Single-nucleotide polymorphisms used as instrumental variables for the Mendelian randomization analysis of GrimAge age acceleration and GGT.** Effect estimates and allele frequencies are shown for the exposure and outcome datasets after harmonization. EA, effect allele; OA, other allele; EAF, effect allele frequency.

**Supplementary Table S20. Single-nucleotide polymorphisms used as instrumental variables for the Mendelian randomization analysis of GrimAge age acceleration and LDL.** Effect estimates and allele frequencies are shown for the exposure and outcome datasets after harmonization. EA, effect allele; OA, other allele; EAF, effect allele frequency.

**Supplementary Table S21. Single-nucleotide polymorphisms used as instrumental variables for the Mendelian randomization analysis of GrimAge age acceleration and WBC.** Effect estimates and allele frequencies are shown for the exposure and outcome datasets after harmonization. EA, effect allele; OA, other allele; EAF, effect allele frequency.

**Supplementary Table S22. Mediation analyses of the associations between environmental exposure biomarkers and GGT through GrimAge age acceleration.** Each log2-transformed exposure biomarker was evaluated separately, with GrimAge age acceleration specified as the mediator and log2-transformed GGT as the outcome. Models were adjusted for age, sex, smoking status, BMI, and estimated blood cell proportions. Average causal mediation effects (ACMEs), average direct effects (ADEs), total effects, and proportions mediated were estimated using 5,000 bootstrap simulations. FDR-adjusted P values for the ACMEs were calculated across the 13 environmental exposure biomarkers.

**Supplementary Table S23. Sensitivity analyses of the associations between environmental exposure biomarkers and GGT through GrimAge age acceleration with additional adjustment for current drinking status.** Each log2-transformed exposure biomarker was evaluated separately, with GrimAge age acceleration specified as the mediator and log2-transformed GGT as the outcome. Models were adjusted for age, sex, smoking status, BMI, estimated blood cell proportions, and current drinking status. Average causal mediation effects (ACMEs), average direct effects (ADEs), total effects, and proportions mediated were estimated using 5,000 bootstrap simulations. FDR-adjusted P values for the ACMEs were calculated across the 13 environmental exposure biomarkers.

**Supplementary Table S24. Mendelian randomization results for the associations between GrimAge age acceleration and 38 disease outcomes.** The inverse-variance weighted (IVW) method was used as the primary estimator. Instrument strength and robustness were assessed using mean F-statistics, I², Cochran’s Q test for heterogeneity, and the MR-PRESSO global test. FDR-adjusted P values were calculated across all evaluated disease outcomes.

**Supplementary Table S25. Nominally significant Mendelian randomization associations between GrimAge age acceleration and disease outcomes.** Shown are disease outcomes with nominal P < 0.05 in the inverse-variance weighted analysis. FDR-adjusted P values were calculated across all evaluated disease outcomes. Instrument strength and robustness were assessed using F-statistics, I², Cochran’s Q test, and the MR-PRESSO global test.

**Supplementary Table S26. Single-nucleotide polymorphisms used as instrumental variables for the Mendelian randomization analysis of GrimAge age acceleration and diabetic nephropathy.** Effect estimates and allele frequencies are shown for the exposure and outcome datasets after harmonization. EA, effect allele; OA, other allele; EAF, effect allele frequency.

**Supplementary Table S27. Single-nucleotide polymorphisms used as instrumental variables for the Mendelian randomization analysis of GrimAge age acceleration and osteoporosis.** Effect estimates and allele frequencies are shown for the exposure and outcome datasets after harmonization. EA, effect allele; OA, other allele; EAF, effect allele frequency.

**Supplementary Table S28. Single-nucleotide polymorphisms used as instrumental variables for the Mendelian randomization analysis of GrimAge age acceleration and cardiac valvular disease.** Effect estimates and allele frequencies are shown for the exposure and outcome datasets after harmonization. EA, effect allele; OA, other allele; EAF, effect allele frequency.

**Supplementary Table S29. Single-nucleotide polymorphisms used as instrumental variables for the Mendelian randomization analysis of GrimAge age acceleration and dilated cardiomyopathy.** Effect estimates and allele frequencies are shown for the exposure and outcome datasets after harmonization. EA, effect allele; OA, other allele; EAF, effect allele frequency.
